# Digital Therapeutic for Hwa-byung Based on Acceptance and Commitment Therapy: A Pilot Feasibility Trial

**DOI:** 10.64898/2026.04.19.26351203

**Authors:** Chan-Young Kwon, Boram Lee, Minjae Kim, Ji-hu Mun, Min-Gyeong Seo, Dohee Yoon

**Affiliations:** Department of Oriental Neuropsychiatry, Dong-Eui University College of Korean Medicine, Busan, Republic of Korea; Anti-Aging Research Center, Dong-Eui University, Busan, Republic of Korea; Research Institute of Korean Medicine, Dong-Eui University, Busan, Republic of Korea; KM Science Research Division, Korea Institute of Oriental Medicine, Daejeon, Republic of Korea; Department of Korean Internal Medicine, Dong-Eui University College of Korean Medicine, Busan, Republic of Korea

**Keywords:** Hwa-byung, digital therapeutics, acceptance and commitment therapy, pilot study, mobile health application

## Abstract

**Background:** Hwa-byung (HB) is a Korean culture-bound syndrome characterised by prolonged suppression of anger and somatic complaints. No evidence-based digital therapeutic (DTx) has been developed for HB. We evaluated the feasibility, user experience (UX), and preliminary clinical effect of an acceptance and commitment therapy (ACT)-based DTx application, Hwa-free, for HB.

**Methods:** Adults aged 19–80 years diagnosed with HB were enrolled in a four-week app-based intervention with assessment at baseline (Week 0), Week 2, Week 4, and Week 8 follow-up. The primary outcome was UX assessed via a 22-item survey at Week 4. Secondary outcomes included HB-related symptom and personality scales, depression, anxiety, anger expression, psychological flexibility, health-related quality of life, and heart rate variability.

**Results:** Of 45 screened, 30 were enrolled and 28 constituted the modified intention-to-treat population. Mean app use was 19.9 ± 7.9 days (71.2% adherence over 28 days). Adverse events were infrequent and unrelated to the intervention. Positive response rates exceeded 80% for video content (items 2–4: 82.8–89.7%), HB self-assessment (86.2%), meditation therapy (86.2%), and in-app guidance (85.7%). Pre–post improvements from baseline to Week 4 were observed in 11 of 18 clinical scales, including HB Symptom Scale (Δ = −9.8, Cohen’s d = −0.92), Beck Depression Inventory-II (Δ = −13.3, d = −1.11), and state anger (Δ = −7.8, d = −0.96). The HB screening-positive rate declined from 100% at baseline to 55.6% at Week 8.

**Conclusions:** Hwa-free demonstrated adequate feasibility, acceptable UX, and preliminary evidence of clinically meaningful improvement in HB-related symptoms. Future randomised controlled trial is warranted.

**Trial registration:** CRIS, KCT0011105

## 1. Introduction

Hwa-byung (HB), literally meaning anger illness, is a Korean culture-bound syndrome first introduced into the psychiatric literature in 1983 (1). It is characterised by the prolonged suppression of anger and resentment, manifesting as a heterogeneous cluster of somatic and psychological symptoms, including chest tightness, a rising or burning sensation, heat in the face or chest, a lump-like sensation in the throat or epigastrium, palpitations, and a deep-seated feeling of injustice (*han*) (2). The condition has been acknowledged by the American Psychiatric Association as a cultural concept of distress (3) and is estimated to affect between 4.2% and 13.3% of the Korean adult population, disproportionately affecting middle-aged women with limited social support (4). However, recently, a high prevalence rate of 36.3% has been reported among the Korean MZ generation, suggesting that this disease persists in modern Korean culture (5).

Current evidence-based treatments for HB include pharmacotherapy, psychotherapy, and Korean medicine (KM) interventions, but access to specialist care remains constrained by geographic, economic, and stigma-related barriers (4-6). Digital therapeutics (DTx), prescription or over-the-counter software delivering evidence-based therapeutic content, represent a scalable solution to these access challenges (7), and smartphone-based interventions have demonstrated efficacy for depression, anxiety, and insomnia in randomised trials (8, 9). However, no DTx has been developed or rigorously evaluated specifically for HB.

Acceptance and commitment therapy (ACT) is a contextual cognitive-behavioural approach that targets experiential avoidance and promotes psychological flexibility through acceptance, defusion, values clarification, and committed action (10). ACT has demonstrated efficacy across a range of emotional and somatic conditions (11), and its theoretical framework aligns well with the psychopathology of HB, in which chronic suppression of anger represents a form of experiential avoidance (2, 4). Hwa-free is an ACT-based DTx application developed specifically for individuals with HB, integrating structured psychoeducational content, behavioural skills training (diaphragmatic breathing, relaxation, meditation), and daily self-monitoring (12).

The present study aimed to: (1) evaluate the feasibility and adherence of Hwa-free over a four-week intervention period; (2) assess user experience (UX) as the primary outcome; and (3) generate preliminary data on clinical effect across HB-related psychological and physiological outcomes to inform the design of a subsequent randomised controlled trial (RCT).

## 2. Methods

### 2.1. Study design and setting

A single-arm, single-center pilot study was conducted at the Korean Medicine Hospital of Dong-eui University, Busan, South Korea. The study was approved by the Institutional Review Board of Dong-eui University Korean Medicine Hospital (DH-2025-15; October 15, 2025) and submitted to the Clinical Research Information Service (CRIS; KCT0011105) on October 21, 2025, prior to the first participant’s enrolment on October 30, 2025. All participants provided written informed consent prior to enrolment. A detailed study protocol has been published elsewhere (12). Reporting follows the CONSORT extension for pilot and feasibility trials, as applicable to single-arm designs (13).

### 2.2. Participants

Eligible participants were adults aged 19–80 years who met diagnostic criteria for HB as confirmed by the Hwa-byung Diagnostic Interview Schedule (HBDIS) (14). Key exclusion criteria included: initiation or substantial modification of any treatment for HB within the preceding four weeks; concurrent participation in an interventional clinical trial; planned initiation of new HB treatment during the study period; unstable serious psychiatric comorbidity (schizophrenia, bipolar disorder) under active management; severe chronic or terminal illness; or inability to comply with study procedures as judged by the investigator. Participants were permitted to continue pre-existing stable treatments throughout the study period; however, initiation of new treatments for HB during the study was not permitted. Participants were recruited via in-hospital posters from October 30, 2025, to January 22, 2026. Target sample size was 30 participants (minimum 25 required based on prior preliminary study for technology acceptance of DTx (15), with 20% attrition allowance).

### 2.3. Intervention

Hwa-free is a smartphone application delivering a structured four-week ACT-based programme (**Figure 1**). A full description of the intervention modules and their theoretical basis is provided in the published protocol (12); briefly, the programme comprises weekly ACT-informed psychoeducational video content, daily diaphragmatic breathing training (with smartphone accelerometry for breath detection), relaxation therapy, meditation exercises, a three-line journaling module, and a HB symptom self-assessment feature. Automated push notifications were delivered at 22:00 daily to encourage engagement. The intervention period was four weeks (28 days); app access was programmatically restricted after Week 4, and participants had no access to the application during the subsequent four-week observation period (Weeks 4–8).

**Figure 1.**
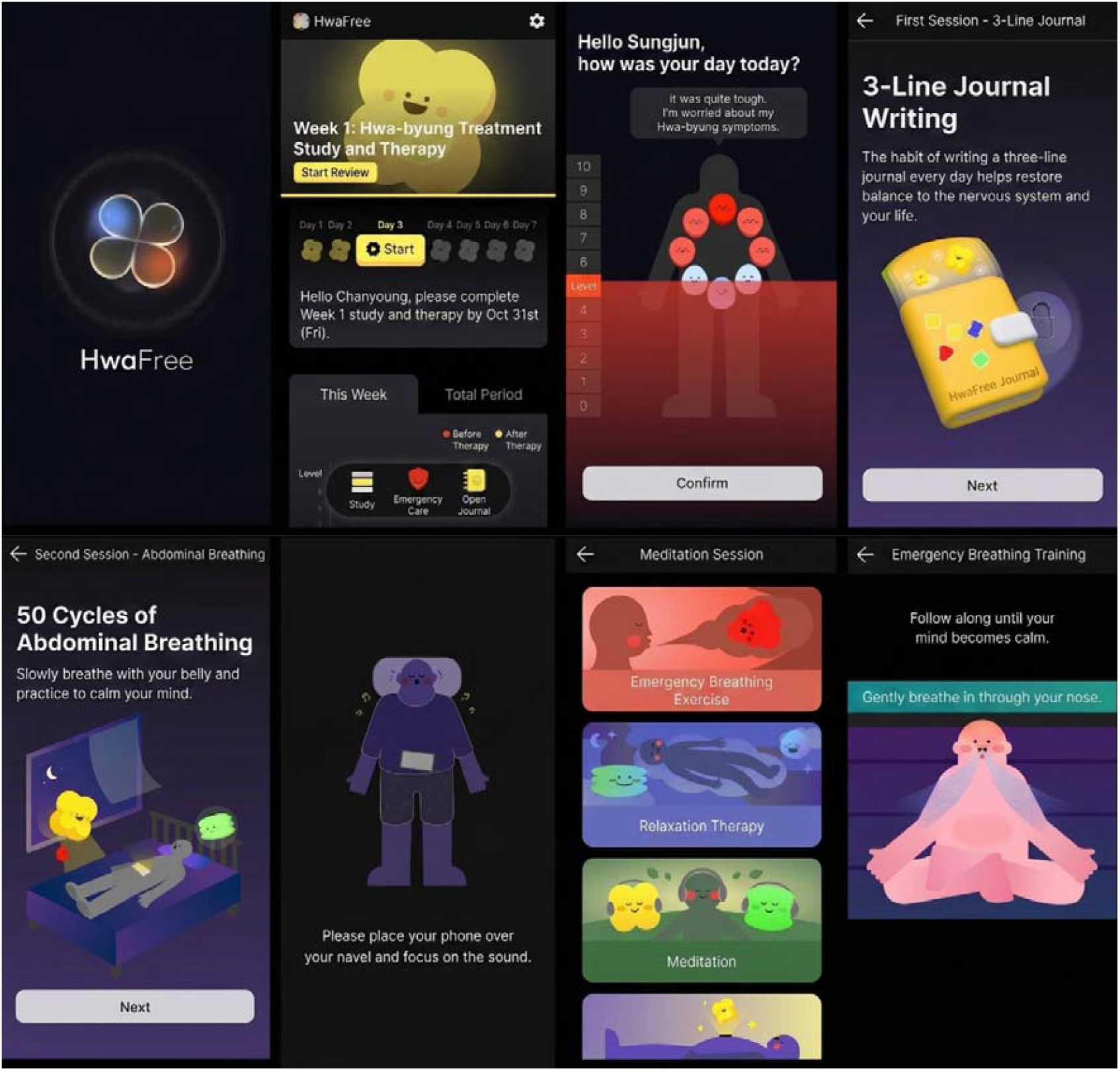
Screenshots of Hwa-free smartphone application interface. **Note**. The screenshots illustrate the comprehensive digital therapeutic components for Hwa-byung management, including: the home interface and treatment progress tracking; daily symptom and stress level assessment; a three-line therapeutic journal; guided abdominal breathing sessions utilizing smartphone-based biofeedback; and emergency relief modules featuring relaxation therapy and guided meditation.

### 2.4. Outcome Measures

#### 2.4.1. Feasibility and adherence

Feasibility was operationalised as daily app use (minimum: app launch; maximum: full diaphragmatic breathing session completion) over the 28-day intervention.

#### 2.4.2. Primary outcome

The primary outcome was UX at Week 4, assessed by a self-developed 22-item survey administered as a self-completed paper questionnaire at the Week 4 clinical visit. The survey was developed iteratively: an initial item pool was generated based on the app’s functional domains, reviewed for content validity by two clinical experts in HB and digital health, and refined following a pilot administration to five individuals with HB experience. Items 1–17 and 20–22 used a 0–4 Likert scale (0 = not at all, 4 = strongly agree); item 16 (barriers to diary use) was reverse-scored. Items 18 and 19 assessed usage frequency (ordinal, 4 categories) and reasons for non-daily use (categorical, multiple-response), respectively; these items were analysed separately and are not included in the domain scoring. Items were grouped into seven a-priori domains: Video Content, Diaphragmatic Breathing, Relaxation Therapy, Meditation Therapy, Three-line Diary, Self-Assessment, and App Usability. Positive response was defined as a score ≥ 3 (or ≤ 1 for item 16). Internal consistency was assessed by Cronbach’s α.

#### 2.4.3 Secondary outcomes

Secondary outcomes included the HB scale including HB Symptom Scale (HBSS; 0–60) and HB Personality Scale (HBPS; 0–64) (16), Beck Depression Inventory-II (BDI-II) (17), State-Trait Anxiety Inventory (STAI; state, STAI-S; trait, STAI-T) (18), State-Trait Anger Expression Inventory (STAXI; state anger, STAXI-S; trait anger, STAXI-T; anger control; anger-out; anger-in) (19), and a 15-item visual analogue scale (VAS; 0–100 mm) for individual HB symptoms. Additional measures included the Acceptance and Action Questionnaire-II (AAQ-II; 8 items, 1–7 Likert scale; higher scores indicate greater experiential avoidance) (20), EQ-5D-5L (Korean utility weights) (21), and heart rate variability (HRV; Canopy9 Plus; 1-min measurement following 5 min of seated rest) (22). HB screening positivity was defined as HBSS ≥ 30 (16). In the absence of an established MCID or validated responder threshold for the HBSS, a ≥ 30% decrease from baseline was adopted as an exploratory responder criterion for descriptive purposes only; this threshold was not pre-specified and findings should be interpreted as hypothesis-generating. Clinical assessments were conducted at Weeks 0, 2, 4, and 8 (or Weeks 0, 4, and 8 for AAQ-II and EQ-5D-5L; Weeks 0 and 4 for HRV).

#### 2.4.4 Safety

Adverse events (AEs) were assessed at Weeks 2 and 4. Investigators recorded the occurrence of AEs and assessed their causal relationship to the intervention using a 6-point causality scale (1 = definitely related; 2 = probably related; 3 = possibly related; 4 = probably not related; 5 = definitely not related; 6 = unknown).

### 2.5. Statistical Analysis

Two analysis populations were defined: (1) the modified intention-to-treat (mITT) population (primary analysis; ≥1 day of app use and ≥1 post-baseline assessment) and (2) the per-protocol (PP) population (sensitivity analysis; Week 4 assessment completed and ≥14 use days). Missing data were handled by complete case analysis for each pairwise comparison; participants with missing data at a given timepoint were excluded only from analyses involving that timepoint, and no imputation was performed. Normality was evaluated by the Shapiro-Wilk test. Pre–post comparisons used paired t-tests (normally distributed difference) or Wilcoxon signed-rank tests (non-normal), with Cohen’s d or r as effect size, respectively. Multiple comparisons were corrected using the Benjamini-Hochberg false discovery rate (FDR). Repeated-measures changes were assessed by Friedman’s test with Dunn’s post-hoc test (Bonferroni-corrected) and mixed models for repeated measures (MMRM; random intercept, baseline as covariate, restricted maximum likelihood). Adherence–outcome associations used Spearman’s ρ. An exploratory mediation analysis examined whether change in AAQ-II (ΔAAQ-II) mediated HBSS change (ΔHBSS), using ordinary least squares regression and bootstrap resampling (N = 5,000; 95% CI); results are hypothesis-generating only given the single-arm design. All analyses were conducted in Python 3.12 (pandas, scipy, statsmodels, scikit-posthocs). Significance threshold: α = 0.05, two-tailed.

## 3. Results

### 3.1. Participants

Of 45 individuals screened for eligibility, 15 were excluded for not meeting the HB diagnostic criteria on the HBDIS and 30 were enrolled (**Figure 2**). The first participant was enrolled on October 30, 2025; the final study assessment was completed on March 31, 2026. Two enrolled participants were excluded from the mITT population: one who withdrew before the Week 2 visit citing a medical reason that precluded smartphone use (no post-baseline clinical assessments available), and one whose app use data were unavailable (due to private VPN settings) and conservatively treated as zero days (mITT: n = 28) (**Table S1**). Nineteen participants met per-protocol criteria (PP: n = 19). Participants were predominantly female (86.7%; n = 26/30), with a median age of 55.5 years (IQR 48.2–65.0) and a median HB duration of 8.5 years (IQR 4.2–20.8). Mean BMI was 24.5 ± 3.0 kg/m^2^. At baseline, all mITT participants (100%) were screen-positive for HB (HBSS ≥ 30; mean HBSS: 46.9 ± 7.8). Mean BDI-II was 38.4 ± 10.4 and mean AAQ-II was 39.7 ± 9.6 (**Table 1**).

**Table 1.** Baseline characteristics of enrolled participants.

| Characteristics | Value (N = 30) |
| --- | --- |
| Age (years)<sup>†</sup> | 55.5 (48.2–65.0) |
| Female, n (%) | 26 (86.7) |
| BMI (kg/m<sup>2</sup>) | 24.48 ± 2.97 |
| HB duration (years) | 8.5 (4.2–20.8) |
| Psychiatric history, n (%) | 3 (10.0) |
| Other medical history, n (%) | 25 (83.3) |
| HBPS | 47.70 ± 9.63 |
| HBSS | 46.93 ± 7.83 |
| BDI-II | 38.37 ± 10.40 |
| STAI-S | 44.97 ± 4.81 |
| STAI-T | 47.90 ± 5.50 |
| STAXI-S | 28.47 ± 7.16 |
| STAXI-T | 28.87 ± 6.88 |
| Anger Control | 19.83 ± 4.24 |
| Anger-Out | 19.00 ± 5.12 |
| Anger-In | 22.90 ± 4.74 |
| AAQ-II | 39.70 ± 9.58 |
| EQ-5D-5L Index | 0.75 (0.59–0.82) |
| EQ-5D-5L VAS | 55.17 ± 18.45 |

**Abbreviations.** AAQ-II, Acceptance and Action Questionnaire-II; BDI-II, Beck Depression Inventory-II; BMI, body mass index; EQ-5D-5L, EuroQol 5-dimension 5-level; HB, Hwa-byung; HBPS, Hwa-byung Personality Scale; HBSS, Hwa-byung Symptom Scale; IQR, interquartile range; SD, standard deviation; STAI-S/-T, State-Trait Anxiety Inventory state/trait subscale; STAXI-S/-T, State-Trait Anger Expression Inventory state/trait subscale. **Note.** Values are presented as n (%), mean ± SD, or median (IQR) based on Shapiro-Wilk test.

**Figure 2.**
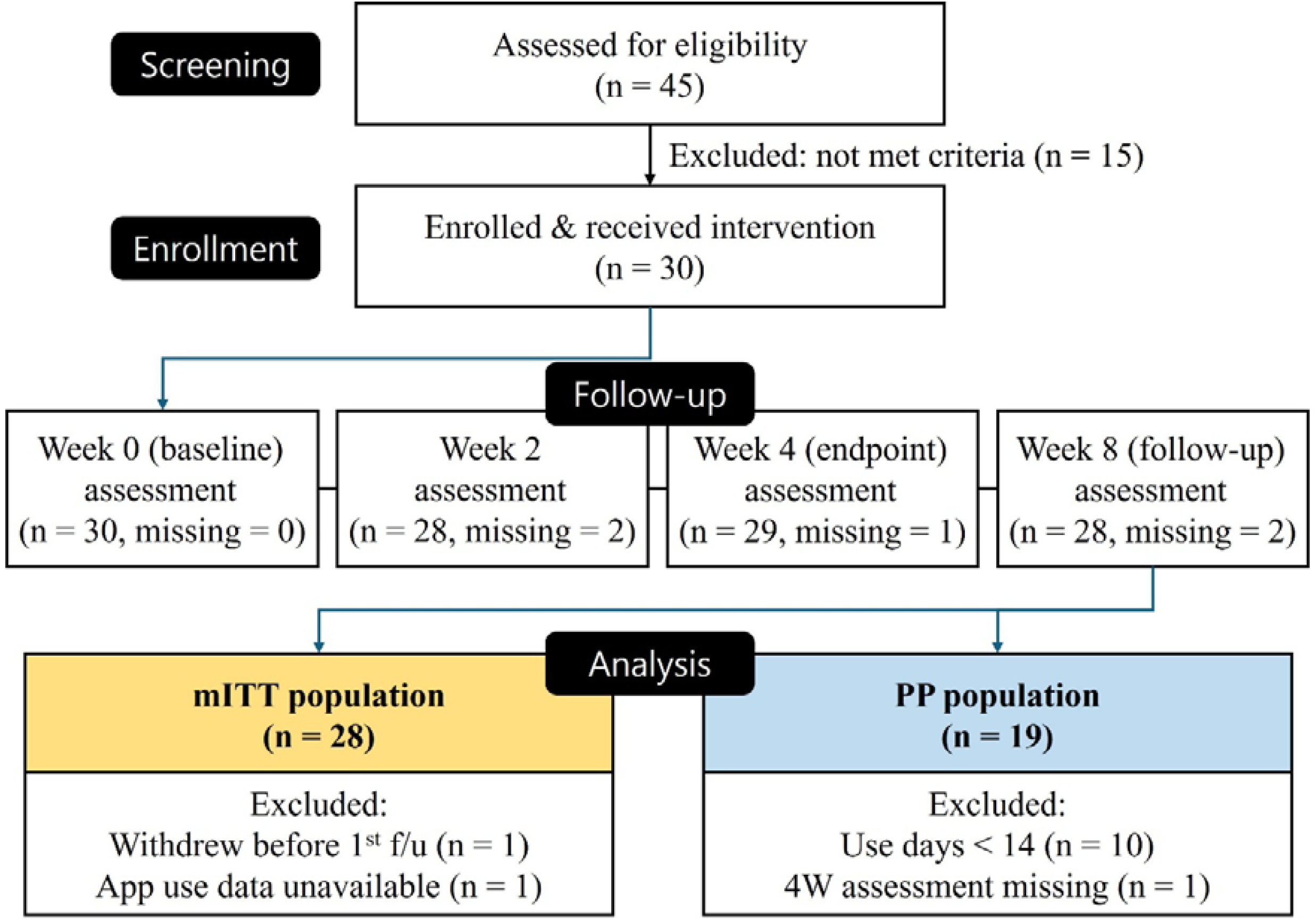
Flow diagram of participant enrollment and follow-up. **Abbreviations**. mITT, modified intention-to-treat; PP, per-protocol. **Note**. All screen failures were attributable to non-fulfilment of HB diagnostic criteria on the HBDIS (n = 15).

### 3.2. Feasibility and Adherence

Participants used the app on a mean of 19.9 ± 7.9 days out of 28 (minimum use-day adherence: 71.2% ± 28.3%). The mean number of days on which the complete diaphragmatic breathing session was performed was 14.4 ± 10.1 (51.5% ± 36.0%) (**Table S2**).

### 3.3. Safety

A total of 8 AEs were reported across the intervention period: 3 events in 2 participants (6.7%) during Weeks 0–2 (insomnia onset, n = 1; chronic tonsillitis and pre-hypertension, n = 1), and 5 events in 3 participants (10.0%) during Weeks 2–4 (hypertension and reflux esophagitis, n = 1; acute nasopharyngitis and allergic dry eye syndrome, n = 1; generalised myalgia, n = 1). All AEs were assessed by the investigator as probably or definitely unrelated to the study intervention, and no serious AEs were reported (**Table S3**).

### 3.4. Primary Outcome: User Experience

Positive response rates (score ≥ 3) were highest for Week 4 video content (item 4: 89.7%), the HB self-assessment function (item 17: 86.2%), meditation therapy effectiveness (item 9: 86.2%), in-app guidance clarity (item 20: 85.7%), and Week 2 video content (item 2: 82.8%). The lowest-rated items were diaphragmatic breathing breath-count accuracy (item 12: 41.4%), ease of placing the phone on the abdomen during breathing (item 11: 48.3%), and ease of following breathing instructions (item 13: 62.1%). The overall internal consistency of the UX questionnaire was Cronbach’s α = 0.9067.

Domain-level internal consistency estimates and item-level score distributions are provided in **Table S4**. Daily app use was reported as daily by 44.8% and 4–5 times per week by 31.0% of respondents (**Table 2**; **Figure S1)**.

**Table 2.**
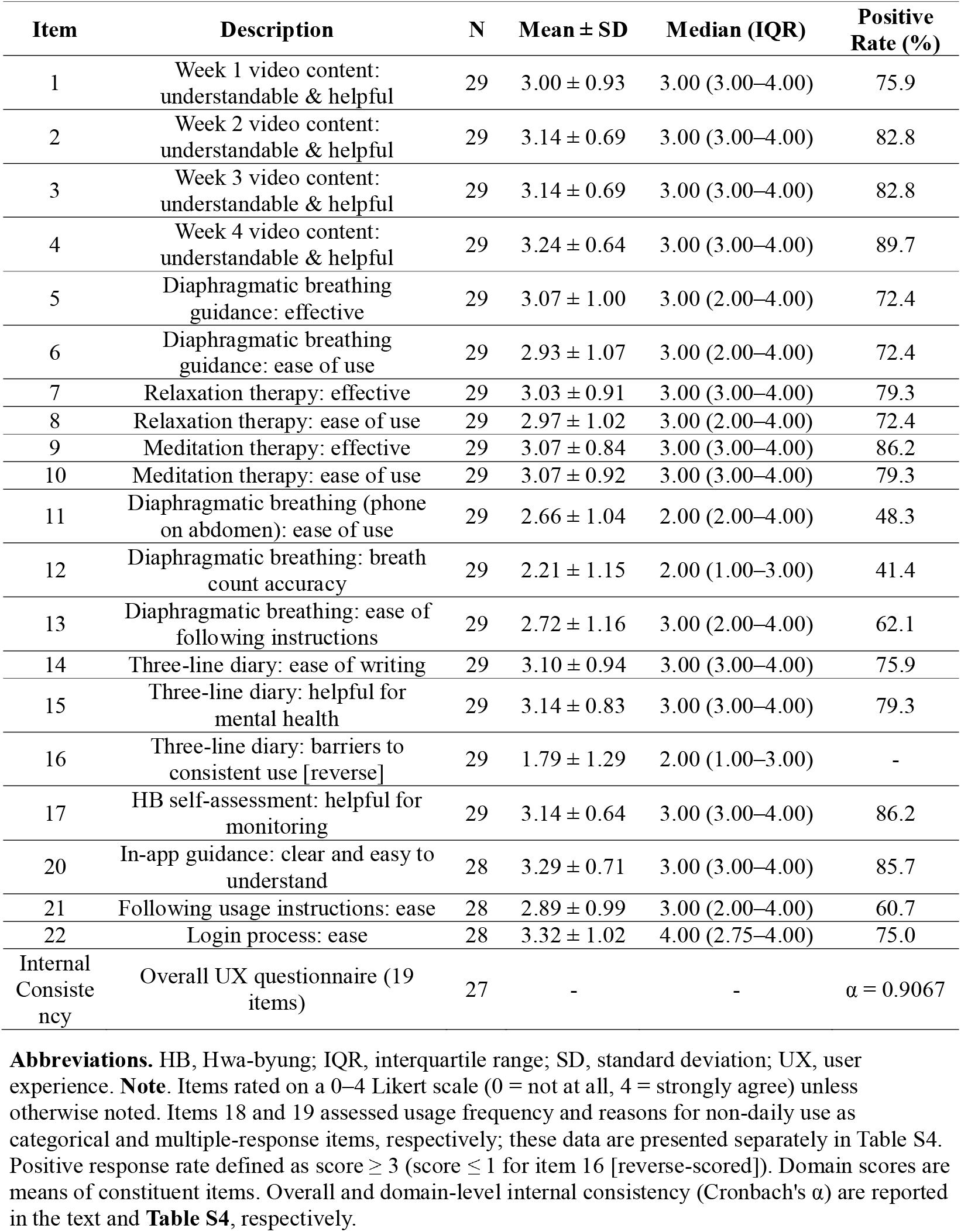
UX survey results at Week 4.

| Item | Description | N | Mean ± SD | Median (IQR) | Positive Rate (%) |
| --- | --- | --- | --- | --- | --- |
| 1 | Week 1 video content: understandable & helpful | 29 | 3.00 ± 0.93 | 3.00 (3.00–4.00) | 75.9 |
| 2 | Week 2 video content: understandable & helpful | 29 | 3.14 ± 0.69 | 3.00 (3.00–4.00) | 82.8 |
| 3 | Week 3 video content: understandable & helpful | 29 | 3.14 ± 0.69 | 3.00 (3.00–4.00) | 82.8 |
| 4 | Week 4 video content: understandable & helpful | 29 | 3.24 ± 0.64 | 3.00 (3.00–4.00) | 89.7 |
| 5 | Diaphragmatic breathing guidance: effective | 29 | 3.07 ± 1.00 | 3.00 (2.00–4.00) | 72.4 |
| 6 | Diaphragmatic breathing guidance: ease of use | 29 | 2.93 ± 1.07 | 3.00 (2.00–4.00) | 72.4 |
| 7 | Relaxation therapy: effective | 29 | 3.03 ± 0.91 | 3.00 (3.00–4.00) | 79.3 |
| 8 | Relaxation therapy: ease of use | 29 | 2.97 ± 1.02 | 3.00 (2.00–4.00) | 72.4 |
| 9 | Meditation therapy: effective | 29 | 3.07 ± 0.84 | 3.00 (3.00–4.00) | 86.2 |
| 10 | Meditation therapy: ease of use | 29 | 3.07 ± 0.92 | 3.00 (3.00–4.00) | 79.3 |
| 11 | Diaphragmatic breathing (phone on abdomen): ease of use | 29 | 2.66 ± 1.04 | 2.00 (2.00–4.00) | 48.3 |
| 12 | Diaphragmatic breathing: breath count accuracy | 29 | 2.21 ± 1.15 | 2.00 (1.00–3.00) | 41.4 |
| 13 | Diaphragmatic breathing: ease of following instructions | 29 | 2.72 ± 1.16 | 3.00 (2.00–4.00) | 62.1 |
| 14 | Three-line diary: ease of writing | 29 | 3.10 ± 0.94 | 3.00 (3.00–4.00) | 75.9 |
| 15 | Three-line diary: helpful for mental health | 29 | 3.14 ± 0.83 | 3.00 (3.00–4.00) | 79.3 |
| 16 | Three-line diary: barriers to consistent use [reverse] | 29 | 1.79 ± 1.29 | 2.00 (1.00–3.00) | - |
| 17 | HB self-assessment: helpful for monitoring | 29 | 3.14 ± 0.64 | 3.00 (3.00–4.00) | 86.2 |
| 20 | In-app guidance: clear and easy to understand | 28 | 3.29 ± 0.71 | 3.00 (3.00–4.00) | 85.7 |
| 21 | Following usage instructions: ease | 28 | 2.89 ± 0.99 | 3.00 (2.00–4.00) | 60.7 |
| 22 | Login process: ease | 28 | 3.32 ± 1.02 | 4.00 (2.75–4.00) | 75.0 |
| Internal Consistency | Overall UX questionnaire (19 items) | 27 | - | - | $\alpha = 0.9067$ |

**Abbreviations.** HB, Hwa-byung; IQR, interquartile range; SD, standard deviation; UX, user experience. **Note.** Items rated on a 0–4 Likert scale (0 = not at all, 4 = strongly agree) unless otherwise noted. Items 18 and 19 assessed usage frequency and reasons for non-daily use as categorical and multiple-response items, respectively; these data are presented separately in Table S4. Positive response rate defined as score $\geq 3$ (score $\leq 1$ for item 16 [reverse-scored]). Domain scores are means of constituent items. Overall and domain-level internal consistency (Cronbach's $\alpha$ ) are reported in the text and **Table S4**, respectively.

### 3.5. Secondary outcomes: Clinical scale changes (Week 4, mITT)

Pre–post comparisons from baseline to Week 4 in the mITT population (n = 28) are summarised in **Table 3** and **Figure 3**. Preliminary improvements were already apparent at Week 2 for several state measures: HBSS declined from 47.1 ± 7.8 at baseline to 41.2 ± 8.6 at Week 2, BDI-II from 38.9 ± 10.4 to 30.6 ± 8.6, STAXI-S from 28.9 ± 7.2 to 24.0 ± 7.9, and STAI-S from 45.0 ± 4.8 to 42.7 ± 5.0.

**Table 3.**
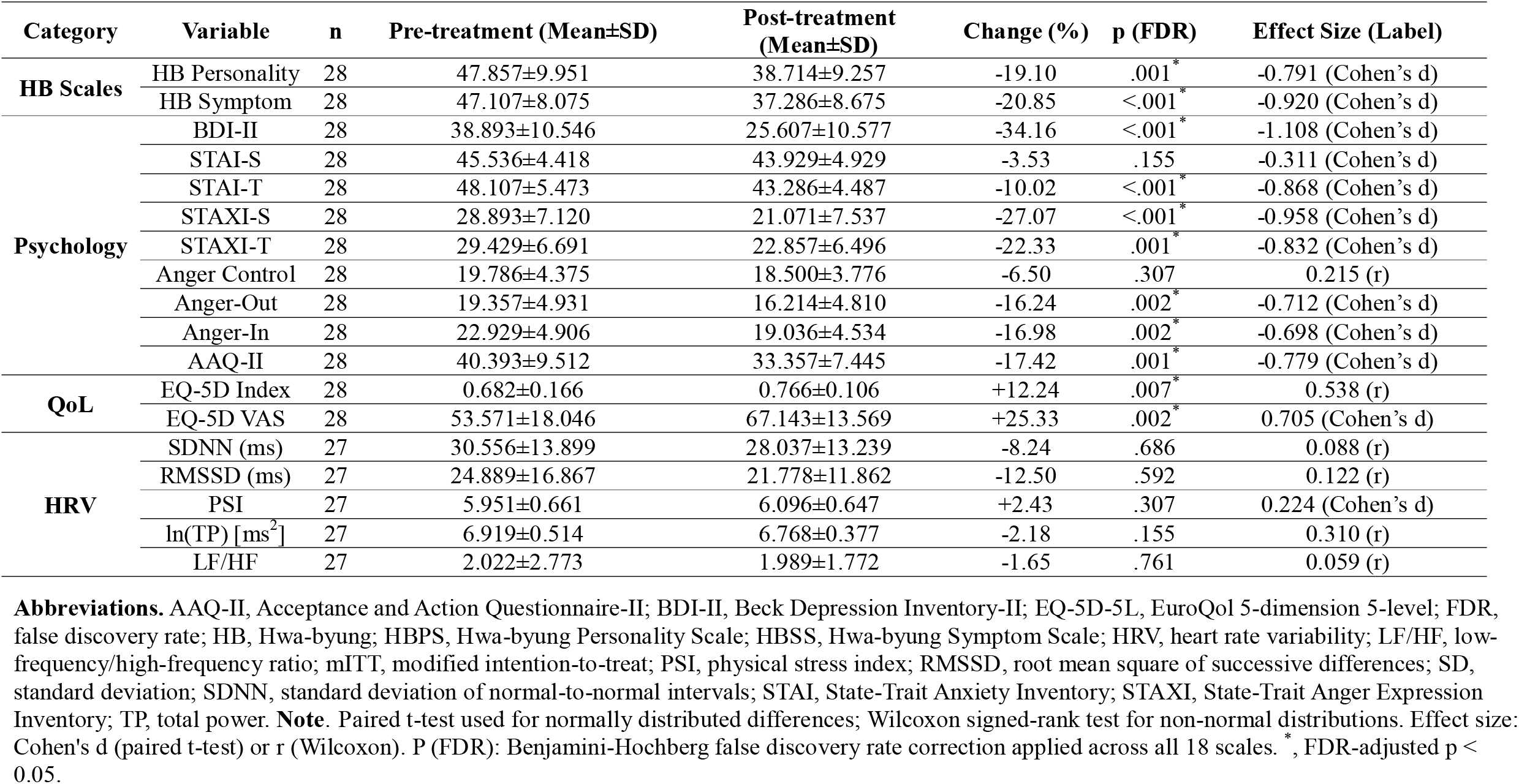
Clinical scale changes from baseline to Week 4 in the mITT population.

**Figure 3.**
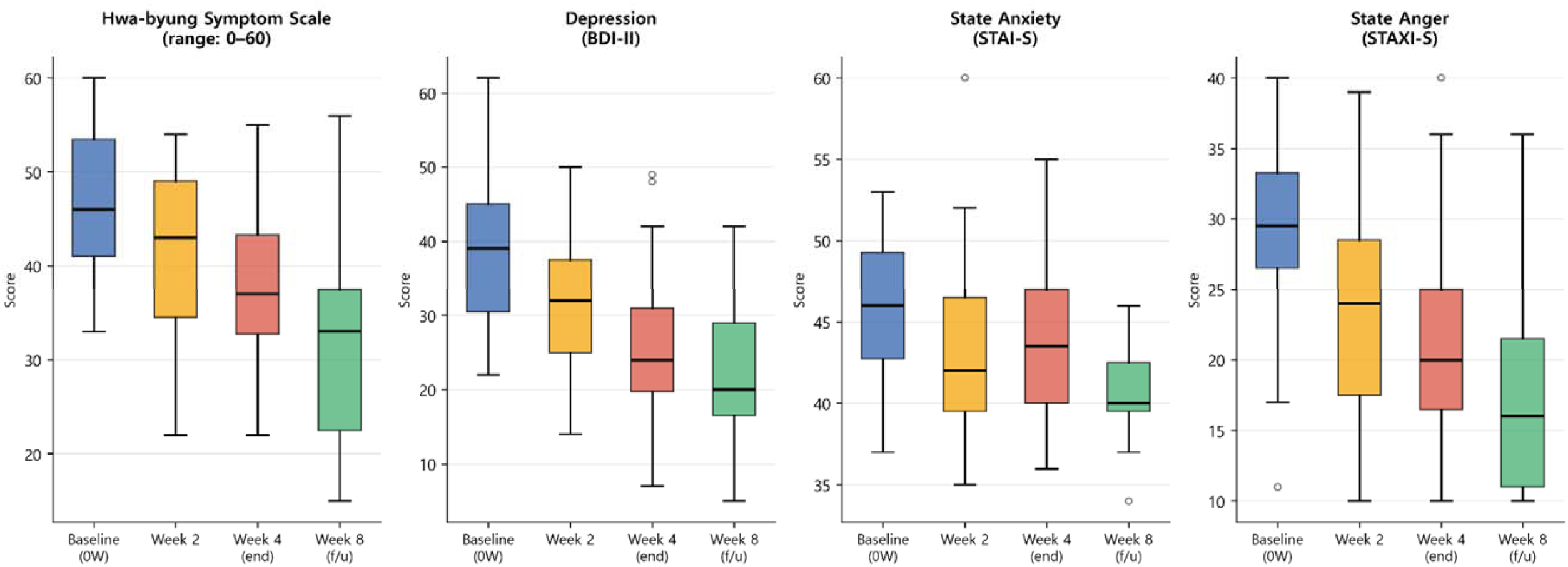
Changes in state/acute clinical outcomes across four timepoints in the mITT population. **Abbreviations**. BDI-II, Beck Depression Inventory-II; STAI-S, State-Trait Anxiety Inventory state subscale; STAXI-S, State-Trait Anger Expression Inventory state anger subscale. **Note**. Box plots show median, interquartile range, and individual paired trajectories (0W to 4W). Colours represent timepoints: baseline (0W, blue), Week 2 (orange), Week 4 (red), Week 8 follow-up (green). Figure 2 presents state and acute symptom measures (Hwa-byung Symptom Scale, Depression [BDI-II], State Anxiety [STAI-S], and State Anger [STAXI-S]); trait- and disposition-level scales (HBPS, STAI-T, STAXI-T, AAQ-II, and others) are presented in Table 3.

Repeated measures post-hoc analysis (Dunn’s test, Bonferroni-corrected) indicated that HBSS and BDI-II did not reach significance at the 2W timepoint (p = 0.187 and p = 0.055, respectively).

Detailed post-hoc results for trait-level scales across all timepoints are presented in **Table S5**.

Of 18 clinical scales analysed, 11 reached statistical significance after FDR correction at Week 4. The largest improvements were observed for BDI-II [Δ = −13.3 (95% CI −17.9 to −8.6), 34.2% reduction, Cohen’s d = −1.11, p < 0.001], STAXI-S [Δ = −7.8 (95% CI −11.0 to −4.7), 27.1% reduction, d = −0.96, p < 0.001], and HBSS [Δ = −9.8 (95% CI −14.0 to −5.7), 20.9% reduction, d = −0.92, p < 0.001]. Significant improvements were also found for HBPS [Δ = −9.1 (95% CI −13.6 to −4.7), d = −0.79], STAI-T [Δ = −4.8 (95% CI −7.0 to −2.7), d = −0.87], STAXI-T [Δ = −6.6 (95% CI −9.6 to −3.5), d = −0.83], AAQ-II [Δ = −7.0 (95% CI −10.5 to −3.5), d = −0.78], Anger-Out [Δ = −3.1 (95% CI −4.9 to −1.4), d = −0.71], Anger-In [Δ = −3.9 (95% CI −6.1 to −1.7), d = −0.70], EQ-5D-5L VAS [Δ = +13.6 (95% CI +6.1 to +21.0), d = 0.71], and EQ-5D-5L Index [Δ = +0.083 (95% CI +0.024 to +0.142), r = 0.54, Wilcoxon]. Fourteen of 15 HB-VAS items showed statistically significant improvement; item 10 (dry mouth) did not reach significance (**Table S6**). HRV indices showed no significant change (all FDR-adjusted p > 0.05; **Figure S2**). PP sensitivity analyses yielded directionally consistent results with comparable effect magnitudes across most significant scales (**Table S7**); however, HB-VAS improvements were substantially attenuated, with only one item (accumulated anger/rage) reaching significance after FDR correction, likely reflecting reduced statistical power in the smaller PP sample (n = 19).

### 3.6. Follow-up outcomes (Week 8, mITT)

Outcomes at Week 8 are presented in **Table S8**. The HB screening-positive rate declined from 100% at baseline to 82.1% at Week 4 and 55.6% at Week 8 (**Table S9**). All 15 HB-VAS items showed significant improvement from baseline to Week 8. Friedman’s test identified significant repeated-measures effects in 27 of 28 scales (**Table S5**). Post-hoc analysis (Dunn’s test, Bonferroni-corrected) indicated that improvements in HBSS and BDI-II were primarily driven by the 0W versus 4W and 0W versus 8W contrasts; the 2W versus 4W and 4W versus 8W contrasts were not significant for either scale, suggesting that change was established by Week 4 and maintained through the app-free observation period. STAI-S reached significance only at the 0W versus 8W contrast (p = 0.0015). Notably, for AAQ-II, the 4W versus 8W contrast was also significant (p = 0.041), indicating that psychological flexibility continued to improve during the app-free observation period. Detailed post-hoc comparisons for all scales are presented in **Table S5**. MMRM confirmed time-related effects in all six primary scales, with all models converging (**Table S10**). HBSS exploratory responder rate (≥ 30% decrease from baseline) increased from 28.6% (8/28) at Week 4 to 59.3% (16/27) at Week 8 (**Table S11**).

### 3.7. Exploratory analyses

Exploratory mediation analyses are presented in **Table S12** and **Figure S3**. ΔAAQ-II was significantly correlated with ΔHBSS (Spearman ρ = 0.53, p = 0.004), and ΔAAQ-II itself showed significant change (Wilcoxon W = 41.5, mean Δ = −7.0, p = 0.001). In the mediation model (controlling for baseline HBSS), ΔAAQ-II significantly predicted ΔHBSS (β = 0.41, p = 0.041); the positive β indicates that greater reduction in AAQ-II scores (i.e., greater improvement in psychological flexibility) was associated with greater reduction in HBSS, consistent with a mediating pathway. The bootstrap indirect effect was significant (a×b = −3.02, 95% CI −6.42 to −0.40, excluding zero; R^2^= 0.46). Adherence (minimum and maximum use days) was not significantly correlated with clinical improvement in any outcome (Spearman’s ρ, all FDR-adjusted p > 0.05; **Table S13**).

## 4. Discussion

The DTx for HB demonstrated adequate feasibility: mean adherence of 71.2% over the 28-day intervention exceeded the upper bound of engagement rates typically reported for mental health smartphone applications in clinical trials (40%–70%) (23, 24), and no participant withdrew due to AEs related to the intervention. Regarding recruitment feasibility, all screen failures were attributable exclusively to HBDIS non-fulfilment (n = 15, 33.3%), suggesting that in-hospital poster recruitment effectively targeted a symptomatic population; future trials may benefit from broader community-based recruitment strategies to improve screening yield. UX was rated favourably across most functional domains, with positive response rates exceeding 72% for video content, relaxation, and meditation modules, supporting the acceptability of these core therapeutic components. UX feedback identified the diaphragmatic breathing module as the primary target for technical refinement: breath-count accuracy (41.4% positive) and the requirement to place the phone on the abdomen (48.3% positive) were the lowest-rated features, and improving the biomechanical sensing interface should be prioritised in future iterations. Among participants who did not use the app daily (n = 21), the most commonly reported barrier was insufficient reminders or alerts (42.9%), followed by content perceived as too long or burdensome (23.8%); only 9.5% cited low perceived immediate need. These findings suggest that optimising notification frequency and timing, and reducing session length or perceived burden, should be prioritised in subsequent iterations.

For contextualisation, the KM Clinical Practice Guideline for HB reports a meta-analytic between-group mean difference of −13.65 points on the HBSS (95% CI −18.24 to −9.05) for face-to-face group ACT versus wait-list control (4). The within-group HBSS change observed in the present study (Δ = −9.8) is directionally consistent with this estimate, although direct comparison is confounded by the absence of a control condition and differences in delivery format and session structure. These findings collectively suggest that ACT-based digital delivery may produce clinically meaningful symptom changes of a broadly comparable magnitude to established face-to-face formats, a hypothesis that warrants confirmation in a head-to-head RCT.

The pattern of improvement is theoretically coherent within the ACT framework: ACT targets experiential avoidance and promotes acceptance of aversive internal states (10), mechanisms directly relevant to the anger suppression that characterises HB (2, 4). Consistent with this, AAQ-II showed significant pre–post improvement (d= −0.78), and the exploratory mediation analysis indicated that ΔAAQ-II was statistically consistent with a partial mediating role in HBSS reduction (indirect effect −3.02, 95% CI −6.42 to −0.40). Of particular interest, significant improvement was also observed in HBPS (d = −0.79), suggesting that the intervention may have engaged dispositional vulnerability factors, habitual emotional suppression and experiential avoidance (16), beyond acute symptom modulation; this hypothesis warrants investigation in future controlled studies with repeated trait-level assessments.

During the four-week app-free observation period (Weeks 4–8), improvement continued across most outcomes: AAQ-II showed a significant further increase between Weeks 4 and 8 (post-hoc p = 0.041), and the exploratory HBSS responder rate doubled from 28.6% at Week 4 to 59.3% at Week 8. This pattern supports internalisation of ACT-based skills acquired during the active programme, rather than dependence on continued app engagement, and is consistent with delayed skill consolidation reported for ACT interventions in other emotional disorders (10). Similar sustained effects have been reported in smartphone-delivered ACTs. In a study by Gentili et al., an 8-week smartphone-delivered ACT intervention was applied to patients with chronic pain, and the effects on psychological flexibility, depression, and anxiety were found to have lasted for more than 6 months (25). In the pathology of HB, the failure to accept or avoid stressful situations is emphasized as a key cause of HB development (2, 4). Therefore, while it is possible that ACT acts on this etiology to contribute to the maintenance of long-term effects, further research is required to clarify the underlying mechanism.

State anxiety (STAI-S) and anger control did not reach statistical significance at Week 4. The STAI-S change at Week 4 (Δ = −1.6) fell substantially below the published anchor-based MCID of approximately 10 points (26), though STAI-S did reach significance at the 0W versus 8W contrast (p = 0.0015), suggesting a possible delayed response. The absence of improvement in anger control is conceptually consistent with the ACT model, which emphasises acceptance over deliberate control of aversive emotions (10); accordingly, anger control was not a primary therapeutic target of the intervention. The absence of significant HRV change likely reflects both the adoption of a one-minute measurement epoch, limiting reliability relative to the five-minute standard (22), and insufficient engagement with the diaphragmatic breathing module, which may be necessary for meaningful autonomic restoration (27). Adherence was not significantly associated with clinical improvement across any outcome, which may reflect a threshold effect whereby the mean dose achieved (71.2%) was sufficient to engage therapeutic mechanisms, with limited additional gain from higher use; alternatively, the small sample may have been underpowered to detect a dose–response relationship.

Several limitations warrant acknowledgement. First, the single-arm design precludes efficacy conclusions: regression to the mean, Hawthorne effects (28), and natural remission cannot be excluded, and effect sizes should not be interpreted as estimates of treatment efficacy or used directly for RCT sample size calculations without appropriate adjustment. Second, no a priori feasibility thresholds were defined; future pilot trials should pre-specify success criteria for adherence and UX. Third, generalisability is restricted by the cultural specificity of HB (1), the predominantly female sample (86.7%), and recruitment from an university affiliated KM hospital. Fourth, the UX survey lacks formal psychometric validation beyond internal consistency. Fifth, missing data (6.7–10% at follow-up) were handled by complete case analysis without sensitivity analyses, limiting the robustness of missing-data conclusions. Sixth, self-reported outcomes, absence of assessor blinding, and unassessed digital literacy introduce potential response and performance bias. Finally, HB-VAS findings should be interpreted with caution, as improvements observed in the mITT analysis were substantially attenuated in the PP sensitivity analysis, suggesting that the current sample may have been underpowered to detect VAS changes in a more conservative analytic framework.

## 5. Conclusions

Hwa-free demonstrated adequate feasibility, acceptable user experience, and preliminary evidence of clinically meaningful improvement across HB symptoms, depression, and anger, with benefits persisting through a four-week app-free follow-up period. These findings support the conduct of a fully powered randomised controlled trial incorporating an active control condition to establish efficacy.

## CRediT authorship contribution statement

**Chan-Young Kwon**: Conceptualization, Methodology, Software, Validation, Formal analysis, Investigation, Data curation, Writing – original draft, Writing – review & editing, Visualization, Supervision, Project administration, Funding acquisition.

**Boram Lee**: Methodology, Writing – review & editing.

**Minjae Kim**: Investigation, Data curation.

**Ji-hu Mun**: Investigation, Data curation.

**Min-Gyeong Seo**: Formal analysis, Data curation.

**Dohee Yoon**: Investigation, Data curation.

## Declaration of competing interest

The authors declare that there is no conflict of interest in the publication of this article.

## Funding

This work was supported by a grant from the Korea Health Technology R&D Project through the Korea Health Industry Development Institute (KHIDI) funded by the Ministry of Health & Welfare, Republic of Korea (grant number: RS-2023-KH139364).

## Ethical statement

The study was approved by the Institutional Review Board of Dong-eui University Korean Medicine Hospital (DH-2025-15; October 15, 2025) and submitted to the Clinical Research Information Service (CRIS; KCT0011105) on October 21, 2025, prior to the first participant’s enrolment on October 30, 2025. All participants provided written informed consent prior to enrolment.

## Data availability

The datasets used and analyzed during the current study are available from the corresponding author on reasonable request.

## Notes

### Competing Interest Statement

The authors have declared no competing interest.

### Clinical Trial

KCT0011105

### Author Declarations

The study was approved by the Institutional Review Board of Dong-eui University Korean Medicine Hospital (DH-2025-15; October 15, 2025).

