## Supplementary Files for "Digital Therapeutic for Hwa-byung Based on Acceptance and Commitment Therapy: A Pilot Feasibility Trial"

**Figure S1. UX survey response distribution at Week 4 (n = 29).**

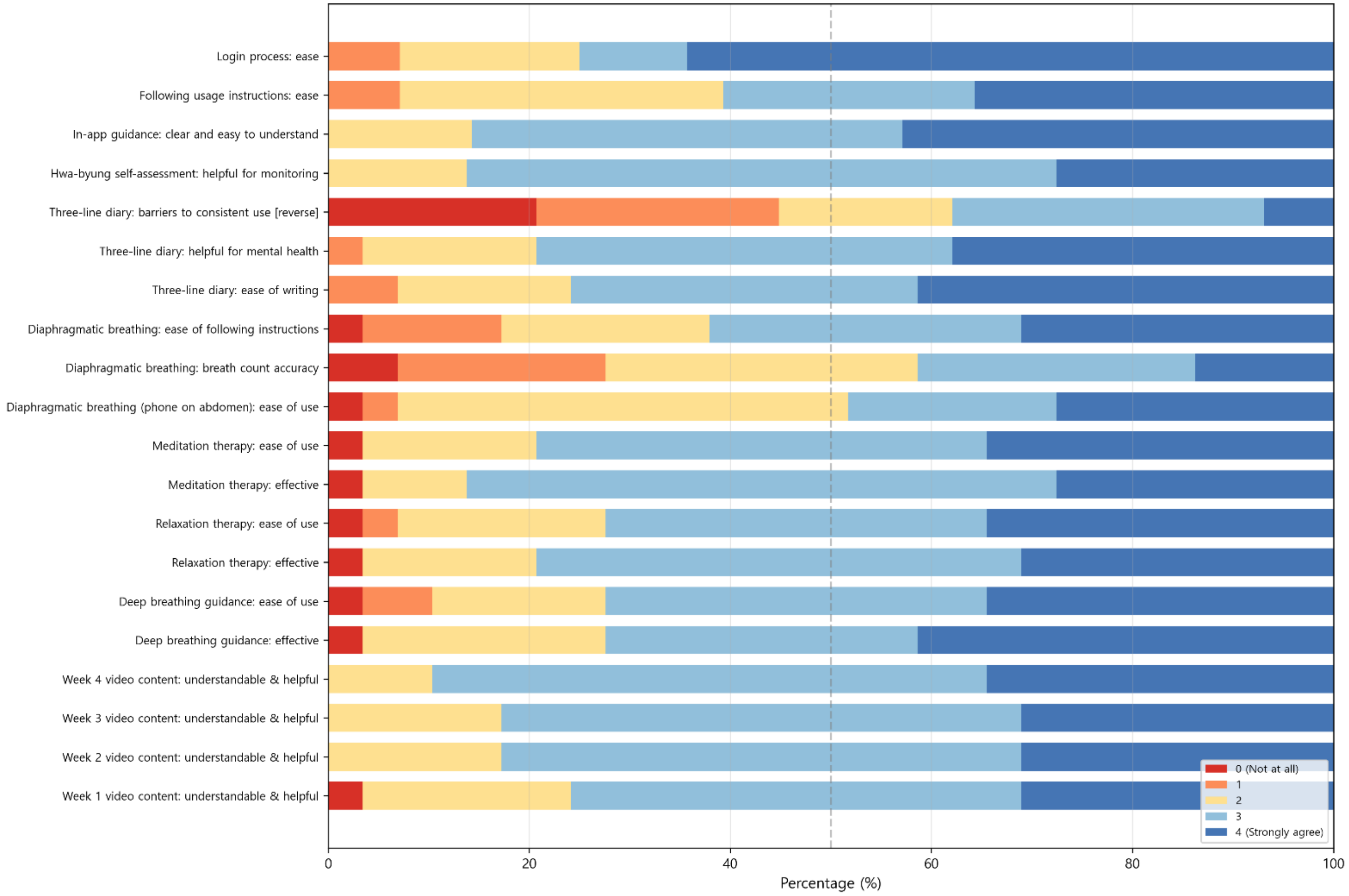


**Note**. Stacked horizontal bar chart showing percentage of respondents selecting each Likert score (0–4) per item. Items ordered as in the questionnaire. Item 16 (reverse-scored) indicated.

**Figure S2. HRV changes from baseline to Week 4 (mITT, n=28).**

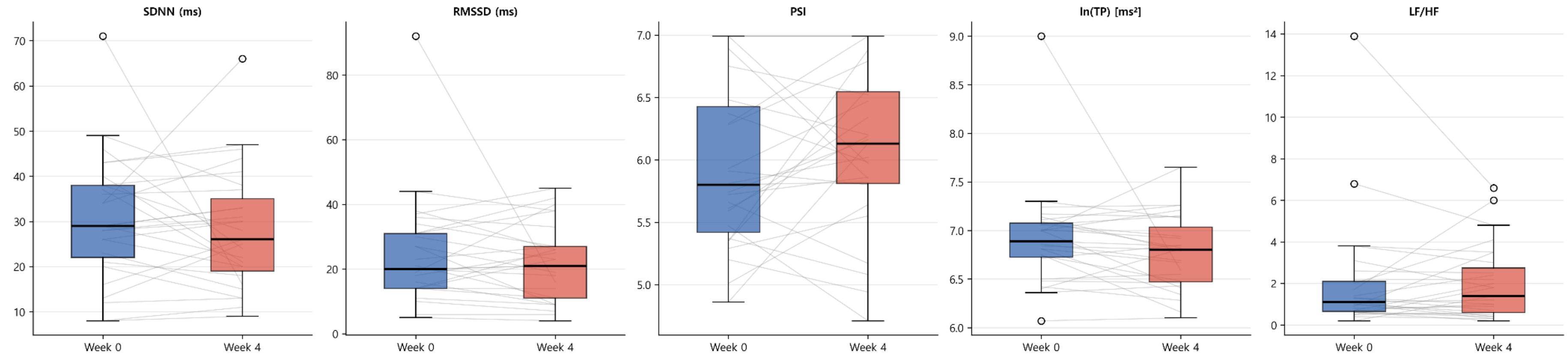


**Note**. Box plots with paired individual trajectories. None of the HRV parameters showed statistically significant change (all FDR-adjusted $p > 0.05$).

**Figure S3. Exploratory ACT-informed mediation analysis.**

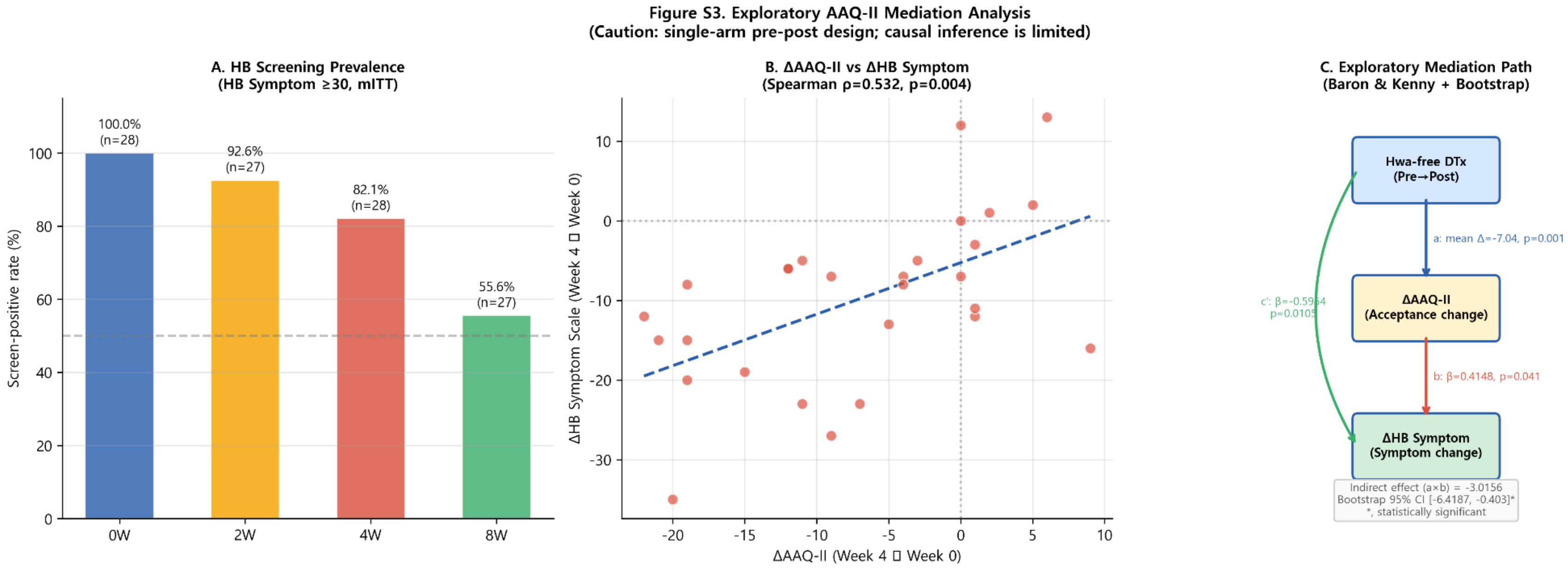


**Note**. Panel A: HB screening-positive rate across timepoints (mITT). Panel B: Scatter plot of ΔAAQ-II versus ΔHBSS with Spearman correlation. Panel C: Mediation path diagram (Baron & Kenny framework with bootstrap). All findings are exploratory; causal inference is not warranted from a single-arm design. ΔAAQ-II = change in Acceptance and Action Questionnaire-II (negative = improvement); ΔHBSS = change in Hwa-byung Symptom Scale (negative = improvement).

**Table S1. Missing data summary by assessment timepoint and key variable.**

| Timepoint | Variable | N of missing | % | N of available |
|---|---|---|---|---|
| **0W** | AAQ-II | 0 | 0 | 30 |
| | Anger-Control | 0 | 0 | 30 |
| | Anger-In | 0 | 0 | 30 |
| | Anger-Out | 0 | 0 | 30 |
| | BDI-II | 0 | 0 | 30 |
| | EQ-5D-5L | 0 | 0 | 30 |
| | HB Personality | 0 | 0 | 30 |
| | HB Symptom | 0 | 0 | 30 |
| | HB-VAS 15 items | 0 | 0 | 30 |
| | HRV | 1 | 3.3 | 29 |
| | STAI-S | 0 | 0 | 30 |
| | STAI-T | 0 | 0 | 30 |
| | STAXI-S | 0 | 0 | 30 |
| | STAXI-T | 0 | 0 | 30 |
| **2W** | AE | 2 | 6.7 | 28 |
| | Anger-Control | 2 | 6.7 | 28 |
| | Anger-In | 2 | 6.7 | 28 |
| | Anger-Out | 2 | 6.7 | 28 |
| | BDI-II | 2 | 6.7 | 28 |
| | HB Personality | 2 | 6.7 | 28 |
| | HB Symptom | 2 | 6.7 | 28 |
| | HB-VAS 15 items | 2 | 6.7 | 28 |
| | STAI-S | 2 | 6.7 | 28 |
| | STAI-T | 2 | 6.7 | 28 |
| | STAXI-S | 2 | 6.7 | 28 |
| | STAXI-T | 2 | 6.7 | 28 |
| **4W** | AAQ-II | 1 | 3.3 | 29 |
| | AE | 1 | 3.3 | 29 |
| | Anger-Control | 1 | 3.3 | 29 |
| | Anger-In | 1 | 3.3 | 29 |
| | Anger-Out | 1 | 3.3 | 29 |
| | BDI-II | 1 | 3.3 | 29 |
| | EQ-5D-5L | 1 | 3.3 | 29 |
| | HB Personality | 1 | 3.3 | 29 |
| | HB Symptom | 1 | 3.3 | 29 |
| | HB-VAS 15 items | 1 | 3.3 | 29 |
| | HRV_PSI | 2 | 6.7 | 28 |

| | | | | |
|---|---|---|---|---|
| | STAI-S | 1 | 3.3 | 29 |
| | STAI-T | 1 | 3.3 | 29 |
| | STAXI-S | 1 | 3.3 | 29 |
| | STAXI-T | 1 | 3.3 | 29 |
| | UX questionnaire | 1 | 3.3 | 29 |
| | Use data (min. use, max. use) | 1 | 3.3 | 29 |
| **8W** | AAQ-II | 2 | 6.7 | 28 |
| | Anger-Control | 2 | 6.7 | 28 |
| | Anger-In | 2 | 6.7 | 28 |
| | Anger-Out | 2 | 6.7 | 28 |
| | BDI-II | 2 | 6.7 | 28 |
| | EQ-5D-5L | 2 | 6.7 | 28 |
| | HB Personality | 2 | 6.7 | 28 |
| | HB Symptom | 2 | 6.7 | 28 |
| | HB-VAS 15 items | 2 | 6.7 | 28 |
| | STAI-S | 2 | 6.7 | 28 |
| | STAI-T | 2 | 6.7 | 28 |
| | STAXI-S | 2 | 6.7 | 28 |
| | STAXI-T | 2 | 6.7 | 28 |

**Table S2. Feasibility and adherence metrics over the 28-day intervention period.**

| Metric | N | Mean±SD | Median (IQR) | Min | Max | Adherence rate (Mean±SD) |
|---|---|---|---|---|---|---|
| **Minimum use days (app launch)** | 29 | 19.93 ± 7.93 | 22.00 (12.00–27.00) | 4 | 28 | 71.2 ± 28.3 |
| **Maximum use days (full session)** | 29 | 14.41 ± 10.09 | 18.00 (3.00–23.00) | 0 | 28 | 51.5 ± 36.0 |
| **Incomplete session days (Min − Max)** | 30 | 5.33 ± 5.09 | 4.50 (1.25–7.75) | 0 | 19 | |

**Note**. Minimum use days: days on which the app was launched. Maximum use days: days on which the complete diaphragmatic breathing session was performed.

**Table S3. Adverse events (AEs) by assessment period and causality assessment.**

| Period | Participants with ≥1 AE, n (%) | Total AE events, n | AE description | Causality |
|---|---|---|---|---|
| **Week 0–2** | 2 (6.7) | 3 | Insomnia onset | Probably not related |
| | | | Chronic tonsillitis | Probably not related |
| | | | Pre-hypertension | Definitely not related |
| **Week 2–4** | 3 (10.0) | 5 | Hypertension | Definitely not related |
| | | | Reflux esophagitis | Definitely not related |
| | | | Acute nasopharyngitis | Definitely not related |
| | | | Allergic dry eye syndrome | Definitely not related |
| | | | Generalised myalgia | Probably not related |

**Note**. n (%) denotes number of participants experiencing at least one AE; total AE events may exceed participant count where one participant reported multiple AEs. AE descriptions are reported as assessed by the clinical investigator; MedDRA coding was not applied in this pilot study. No serious AEs were reported. Causality assessed per ICH E2A guidelines.

**Table S4. UX domain scores, internal consistency, and usage patterns.**

Part A. UX Domain Scores and Internal Consistency (N = 29)

| Domain | Items (n) | Mean ± SD | Median (IQR) | Cronbach's α | Interpretation |
|---|---|---|---|---|---|
| **Video Content** | 1–4 (4) | 3.13 ± 0.63 | 3.00 (2.75–3.75) | 0.8706 | Good |
| **Diaphragmatic Breathing** | 5, 6, 11–13 (5) | 2.72 ± 0.95 | 2.60 (2.20–3.40) | 0.9235 | Excellent |
| **Relaxation Therapy** | 7, 8 (2) | 3.00 ± 0.94 | 3.00 (2.50–4.00) | 0.9402 | Excellent |
| **Meditation Therapy** | 9, 10 (2) | 3.07 ± 0.82 | 3.00 (3.00–3.50) | 0.8410 | Good |
| **Three-line Diary** | 14, 15 (2) | 3.12 ± 0.78 | 3.00 (3.00–4.00) | 0.6885 | Questionable |
| **Self-Assessment** | 17 (1) | 3.14 ± 0.64 | 3.00 (3.00–4.00) | - | - |
| **App Usability*** | 20–22 (3) | 3.13 ± 0.79 | 3.33 (2.33–4.00) | 0.7835 | Acceptable |

**Note**. *, n = 27 due to missing data on item 22.

Part B. Usage Frequency and Reasons for Non-daily Use

| Usage Frequency (Item 18) | n (%) | Reasons for non-daily use (Item 19)† | n (% of respondents) |
|---|---|---|---|
| Daily | 13 (44.8) | Insufficient reminders/alerts | 9 (42.9) |
| 4–5 times/week | 9 (31.0) | Content too long or burdensome | 5 (23.8) |
| 2–3 times/week | 5 (17.2) | Did not feel immediate need | 2 (9.5) |
| Rarely | 2 (6.9) | Difficult to access features | 2 (9.5) |
| | | Other | 6 (28.6) |

**Note**. †, Multiple responses allowed (N = 21 respondents who did not use the app daily).

**Table S5. Repeated measures analysis: Friedman test and Dunn's post-hoc comparisons.**

Part A. mITT Analysis (n = 26–27)

| Category | Variable | χ2 | p-value | 0W (Mean±SD) | 2W (Mean±SD) | 4W (Mean±SD) | 8W (Mean±SD) | Post-hoc Sig. (Dunn) |
|---|---|---|---|---|---|---|---|---|
| **HB Scales** | HB Personality | 41.44 | <.001 | 48.58±9.68 | 40.81±9.72 | 37.54±8.43 | 35.23±7.19 | 0W>4W, 8W |
| | HB Symptom | 35.44 | <.001 | 47.73±8.03 | 41.19±8.58 | 36.77±8.63 | 31.08±10.51 | 0W>4W, 8W; 2W>8W |
| **Psychology** | BDI-II | 36.13 | <.001 | 39.92±10.18 | 30.58±8.56 | 25.23±10.49 | 22.31±9.95 | 0W>4W, 8W; 2W>8W |
| | STAI-S | 17.09 | <.001 | 45.42±4.57 | 42.65±5.00 | 43.27±4.41 | 40.81±2.70 | 0W>8W |
| | STAI-T | 26.18 | <.001 | 48.23±5.62 | 42.65±4.60 | 42.92±4.42 | 41.50±5.09 | 0W>2W, 4W, 8W |
| | STAXI-S | 35.76 | <.001 | 29.50±6.94 | 24.04±7.90 | 21.12±7.25 | 17.00±7.18 | 0W>4W, 8W; 2W>8W |
| | STAXI-T | 30.23 | <.001 | 29.92±6.39 | 24.35±5.75 | 22.58±6.32 | 20.50±6.53 | 0W>2W, 4W, 8W |
| | Anger Control | 5.76 | .124 | 19.42±4.27 | 18.38±3.44 | 18.38±3.81 | 17.62±3.15 | None |
| | Anger-Out | 20.80 | <.001 | 19.62±5.02 | 17.46±4.96 | 16.35±4.81 | 15.15±4.23 | 0W>8W |
| | Anger-In | 27.20 | <.001 | 23.15±4.99 | 19.88±4.22 | 18.85±3.59 | 17.19±3.72 | 0W>4W, 8W |
| | AAQ-II | 27.08 | <.001 | 41.19±8.70 | - | 33.85±7.10 | 27.81±7.30 | 0W>4W, 8W; 4W>8W |
| **QoL** | EQ-5D Index | 16.64 | <.001 | 0.68±0.17 | - | 0.77±0.11 | 0.82±0.09 | 0W<8W |
| | EQ-5D VAS | 12.02 | .003 | 54.07±18.19 | - | 66.30±13.05 | 72.37±10.79 | 0W<8W |

**Note**. Within the modified intention-to-treat (mITT) population (n=28), the sample size for the Friedman test ranged from 26 to 27 due to the requirement for complete data across all analyzed time points. For variables analyzed at three time points (0W, 4W, and 8W), such as AAQ-II and EQ-5D, one participant was excluded due to missing data at the 8W follow-up, resulting in n=27. For other clinical scales requiring assessments at four time points (0W, 2W, 4W, and 8W), two participants were excluded due to missing data at either the 2W or 8W visits, resulting in n=26. This discrepancy reflects the overlap of missing assessments at 2W and 8W among the mITT participants.

Part B. mITT Analysis: HB-VAS 15 Items (n = 26)

| No | Symptom | χ2 | p-value | 0W (Mean±SD) | 2W (Mean±SD) | 4W (Mean±SD) | 8W (Mean±SD) | Post-hoc Sig. (Dunn) |
|---|---|---|---|---|---|---|---|---|
| **01** | **Chest tightness/breathlessness** | 22.58 | <.001 | 74.08 ± 19.74 | 68.35 ± 19.45 | 60.77 ± 22.08 | 57.00 ± 22.01 | 0W>8W |
| **02** | **Rising sensation** | 28.26 | <.001 | 76.65 ± 18.14 | 72.00 ± 17.43 | 63.12 ± 18.85 | 56.58 ± 23.21 | 0W>4W, 8W |
| **03** | **Heat sensation (face/chest)** | 27.73 | <.001 | 75.19 ± 19.81 | 70.27 ± 19.72 | 60.42 ± 23.86 | 50.69 ± 25.92 | 0W>8W; 2W>8W |
| **04** | **Lump sensation (throat/epigastrium)** | 29.56 | <.001 | 77.96 ± 21.22 | 68.15 ± 23.72 | 60.23 ± 20.62 | 52.31 ± 24.96 | 0W>4W, 8W |
| **05** | **Feeling of injustice/resentment** | 24.41 | <.001 | 78.27 ± 23.54 | 70.85 ± 22.36 | 61.54 ± 21.85 | 54.58 ± 25.20 | 0W>4W, 8W |

| | | | | | | | | |
|---|---|---|---|---|---|---|---|---|
| **06** | **Accumulated anger/rage** | 30.43 | <.001 | 83.77 ± 18.43 | 74.88 ± 18.71 | 66.73 ± 21.54 | 57.27 ± 24.21 | 0W>4W, 8W |
| **07** | **Palpitations** | 23.23 | <.001 | 74.12 ± 22.24 | 67.12 ± 16.56 | 60.50 ± 20.60 | 49.35 ± 26.78 | 0W>8W |
| **08** | **Sleep disturbance** | 17.50 | .001 | 84.77 ± 18.33 | 77.12 ± 19.03 | 68.12 ± 20.40 | 67.46 ± 21.76 | 0W>4W, 8W |
| **09** | **Headache/dizziness** | 34.13 | <.001 | 76.38 ± 22.66 | 65.73 ± 20.31 | 56.81 ± 21.31 | 47.54 ± 25.51 | 0W>4W, 8W |
| **10** | **Dry mouth** | 23.05 | <.001 | 73.00 ± 25.19 | 64.19 ± 20.85 | 58.83 ± 24.38 | 49.12 ± 26.01 | 0W>8W |
| **11** | **Loss of appetite** | 19.63 | <.001 | 60.38 ± 26.04 | 50.38 ± 23.75 | 45.87 ± 23.25 | 37.58 ± 26.25 | 0W>8W |
| **12** | **Fearfulness/startle response** | 27.25 | <.001 | 75.00 ± 23.52 | 69.42 ± 21.58 | 58.73 ± 22.85 | 50.54 ± 29.38 | 0W>8W |
| **13** | **Intrusive thoughts** | 25.76 | <.001 | 85.58 ± 16.19 | 80.77 ± 16.04 | 72.42 ± 17.32 | 61.58 ± 26.84 | 0W>4W, 8W; 2W>8W |
| **14** | **Frequent sighing** | 29.92 | <.001 | 84.00 ± 15.29 | 75.08 ± 21.20 | 68.85 ± 20.88 | 54.58 ± 26.92 | 0W>4W, 8W; 2W>8W |
| **15** | **Deep-seated resentment (Han)** | 23.82 | <.001 | 81.85 ± 20.77 | 75.27 ± 19.21 | 63.38 ± 23.66 | 54.88 ± 27.19 | 0W>4W, 8W; 2W>8W |

**Note**. Within the modified intention-to-treat (mITT) population (n=28), the sample size for the Friedman test was n=26 for the HB-VAS 15 items requiring assessments at four time points (0W, 2W, 4W, and 8W). Two participants were excluded because they failed to provide complete data at either the 2W or 8W visits, reflecting the overlap of missing assessments within the mITT group. Post-hoc comparisons were performed using Dunn's test with Bonferroni correction.

Part C. PP Analysis (Sensitivity, n = 17–18)

| **Category** | **Variable** | **χ2** | **p-value** | **0W (Mean±SD)** | **2W (Mean±SD)** | **4W (Mean±SD)** | **8W (Mean±SD)** | **Post-hoc Sig. (Dunn)** |
|---|---|---|---|---|---|---|---|---|
| **HB Scales** | HB Personality | 24.94 | <.001 | 47.71±8.36 | 39.18±8.97 | 37.53±8.52 | 34.47±7.88 | 0W>4W, 8W |
| | HB Symptom | 22.15 | <.001 | 47.06±7.55 | 40.41±8.56 | 35.71±9.66 | 28.71±11.22 | 0W>4W, 8W; 2W>8W |
| **Psychology** | BDI-II | 20.36 | <.001 | 39.35±10.64 | 30.00±9.53 | 23.76±12.43 | 21.00±10.56 | 0W>4W, 8W |
| | STAI-S | 8.47 | .037 | 45.53±5.35 | 42.00±4.49 | 42.12±3.89 | 40.71±3.04 | 0W>8W |
| | STAI-T | 16.22 | .001 | 48.00±5.99 | 42.29±4.50 | 42.29±4.67 | 41.35±5.33 | 0W>8W |
| | STAXI-S | 16.37 | .001 | 28.53±7.73 | 22.94±8.75 | 20.94±8.07 | 17.47±7.71 | 0W>8W |
| | STAXI-T | 16.35 | .001 | 28.71±6.92 | 23.00±5.83 | 21.35±7.13 | 20.29±7.03 | 0W>4W, 8W |
| | Anger Control | 3.84 | .279 | 19.29±4.55 | 18.82±3.28 | 18.12±4.26 | 18.24±3.42 | None |
| | Anger-Out | 8.75 | .033 | 18.41±5.03 | 16.41±4.51 | 15.71±5.22 | 14.82±4.57 | None |
| | Anger-In | 11.07 | .011 | 22.35±5.31 | 19.35±3.95 | 18.94±4.19 | 17.47±4.33 | 0W>8W |
| | AAQ-II | 17.94 | <.001 | 40.17±9.75 | - | 33.00±7.79 | 26.61±7.21 | 0W>8W |
| **QoL** | EQ-5D Index | 12.45 | .002 | 0.68±0.18 | - | 0.77±0.10 | 0.83±0.11 | 0W<8W |

| | | | | | | | |
|---|---|---|---|---|---|---|---|
| EQ-5D VAS | 10.39 | .006 | 51.94±16.28 | - | 62.50±12.28 | 72.78±10.32 | 0W<8W |

**Note**. In the per-protocol (PP) population (n=19), the sample size for the Friedman test varied between n=17 and n=18 due to the requirement for complete data across all analyzed time points. For variables measured at three time points (0W, 4W, and 8W), such as AAQ-II and EQ-5D, one participant was excluded due to a missing 8W follow-up visit, resulting in n=18. For clinical scales measured at four time points (0W, 2W, 4W, and 8W), an additional participant who missed the 2W assessment was excluded along with a participant, resulting in n=17. This individual was one of the two participants with missing 2W data reported in the attrition analysis (**Table S1**) who otherwise met the PP criteria.

Part D. PP Analysis: HB-VAS 15 Items (Sensitivity, n = 17)

| No | Symptom | χ2 | p-value | 0W (Mean±SD) | 2W (Mean±SD) | 4W (Mean±SD) | 8W (Mean±SD) | Post-hoc Sig. (Dunn) |
|---|---|---|---|---|---|---|---|---|
| **01** | **Chest tightness/breathlessness** | 12.23 | .007 | 70.88±18.56 | 63.82±16.91 | 56.00±20.90 | 51.76±22.84 | 0W > 8W |
| **02** | **Rising sensation** | 17.67 | <.001 | 73.65±18.89 | 69.12±17.16 | 60.76±19.09 | 50.88±24.64 | 0W > 8W |
| **03** | **Heat sensation (face/chest)** | 15.81 | .001 | 71.71±21.20 | 67.94±19.69 | 59.00±22.53 | 47.53±24.62 | 0W > 8W |
| **04** | **Lump sensation (throat/epigastrium)** | 18.98 | <.001 | 77.12±20.87 | 66.18±21.76 | 59.29±17.82 | 47.71±24.11 | 0W > 8W |
| **05** | **Feeling of injustice/resentment** | 17.34 | .001 | 75.18±26.10 | 69.12±22.10 | 59.82±24.15 | 49.71±25.03 | 0W > 8W |
| **06** | **Accumulated anger/rage** | 18.00 | <.001 | 82.41±19.56 | 72.65±18.72 | 63.35±22.76 | 53.53±25.17 | 0W > 8W |
| **07** | **Palpitations** | 18.51 | <.001 | 69.53±24.15 | 63.53±16.37 | 53.53±19.59 | 44.06±27.57 | 0W > 8W |
| **08** | **Sleep disturbance** | 12.40 | .006 | 79.59±20.18 | 70.29±19.40 | 60.18±18.99 | 60.47±21.05 | 0W > 4W |
| **09** | **Headache/dizziness** | 27.86 | <.001 | 76.88±21.44 | 62.35±20.16 | 53.82±20.58 | 44.12±21.88 | 0W > 4W, 8W |
| **10** | **Dry mouth** | 13.40 | .004 | 74.35±23.79 | 62.65±19.21 | 56.44±23.09 | 43.35±22.79 | 0W > 8W |
| **11** | **Loss of appetite** | 11.61 | .009 | 59.71±24.01 | 50.29±22.18 | 45.44±20.92 | 35.29±26.31 | 0W > 8W |
| **12** | **Fearfulness/startle response** | 17.91 | <.001 | 72.12±23.51 | 65.94±19.77 | 54.53±20.83 | 39.65±26.11 | 0W > 8W; 2W > 8W |
| **13** | **Intrusive thoughts** | 17.28 | .001 | 83.82±15.75 | 78.35±15.47 | 69.47±16.29 | 58.82±22.81 | 0W > 8W; 2W > 8W |
| **14** | **Frequent sighing** | 14.98 | .002 | 82.47±14.89 | 71.59±23.04 | 64.53±22.60 | 51.47±24.99 | 0W > 8W |
| **15** | **Deep-seated resentment (Han)** | 15.91 | .001 | 80.65±22.69 | 73.94±19.76 | 59.47±25.21 | 50.53±27.21 | 0W > 8W |

**Note**. In the per-protocol (PP) population (n=19), the sample size for the Friedman test was n=17 for the HB-VAS 15 items requiring assessments across four time points (0W, 2W, 4W, and 8W). Exclusions included one participant due to a missing 8W follow-up visit and one additional participant due to a missing 2W assessment, resulting in a total of 17 participants with complete longitudinal data. This specific exclusion of the 2W-missing individual corresponds to the attrition details reported in **Table S1**. Post-hoc comparisons were conducted using Dunn's test with Bonferroni correction.

**Table S6. Changes in HB-VAS 15-item scores from baseline to Week 4.**

Part A. mITT Analysis (Primary, n = 28)

| No | Symptom | Baseline | Week 4 | Change | p-value (FDR) | Cohen's d |
|---|---|---|---|---|---|---|
| **01** | **Chest tightness/breathlessness** | 73.07 ± 19.54 | 61.96 ± 21.71 | -11.11 ± 15.04 | 0.0045 | -0.74 |
| **02** | **Rising sensation** | 76.18 ± 18.37 | 64.50 ± 18.95 | -11.68 ± 17.29 | 0.0045 | -0.68 |
| **03** | **Heat sensation (face/chest)** | 75.18 ± 20.24 | 62.54 ± 24.38 | -12.64 ± 23.68 | 0.0150 | -0.53 |
| **04** | **Lump sensation (throat/epigastrium)** | 77.39 ± 21.23 | 62.71 ± 21.88 | -14.68 ± 21.49 | 0.0045 | -0.68 |
| **05** | **Feeling of injustice/resentment** | 76.25 ± 23.83 | 63.93 ± 22.82 | -12.32 ± 27.40 | 0.0264 | -0.45 |
| **06** | **Accumulated anger/rage** | 82.96 ± 17.99 | 67.86 ± 21.23 | -15.11 ± 17.00 | 0.0015 | -0.89 |
| **07** | **Palpitations** | 74.36 ± 21.43 | 61.18 ± 20.71 | -13.18 ± 23.87 | 0.0150 | -0.55 |
| **08** | **Sleep disturbance** | 84.07 ± 19.08 | 70.04 ± 20.90 | -14.04 ± 23.58 | 0.0100 | -0.60 |
| **09** | **Headache/dizziness** | 73.79 ± 23.96 | 59.00 ± 22.29 | -14.79 ± 29.10 | 0.0156 | -0.51 |
| **10** | **Dry mouth** | 71.36 ± 24.98 | 60.52 ± 24.35 | -10.84 ± 29.80 | 0.0649 | -0.36 |
| **11** | **Loss of appetite** | 60.00 ± 25.13 | 46.88 ± 22.84 | -13.13 ± 19.61 | 0.0045 | -0.67 |
| **12** | **Fearfulness/startle response** | 73.21 ± 23.57 | 60.79 ± 23.50 | -12.43 ± 25.38 | 0.0175 | -0.49 |
| **13** | **Intrusive thoughts** | 84.82 ± 17.22 | 73.68 ± 17.51 | -11.14 ± 20.94 | 0.0150 | -0.53 |
| **14** | **Frequent sighing** | 82.46 ± 16.12 | 70.71 ± 21.28 | -11.75 ± 22.86 | 0.0156 | -0.51 |
| **15** | **Deep-seated resentment (Han)** | 79.21 ± 22.25 | 64.75 ± 23.40 | -14.46 ± 28.58 | 0.0156 | -0.51 |

**Note.** Values are presented as mean ± SD. p-values were calculated using paired t-tests.

Part B. PP Analysis (Sensitivity, n = 19)

| No | Symptom | Baseline | Week 4 | Change | p-value (FDR) | Cohen's d |
|---|---|---|---|---|---|---|
| **01** | **Chest tightness/breathlessness** | 69.74 ± 18.14 | 58.26 ± 20.86 | -11.47 ± 17.23 | 0.0647 | -0.67 |
| **02** | **Rising sensation** | 73.26 ± 19.05 | 63.05 ± 19.42 | -10.21 ± 19.00 | 0.0660 | -0.54 |
| **03** | **Heat sensation (face/chest)** | 72.05 ± 21.68 | 62.26 ± 23.62 | -9.79 ± 24.43 | 0.1047 | -0.40 |
| **04** | **Lump sensation (throat/epigastrium)** | 76.37 ± 20.90 | 63.05 ± 20.29 | -13.32 ± 23.51 | 0.0647 | -0.57 |
| **05** | **Feeling of injustice/resentment** | 72.53 ± 25.86 | 63.53 ± 25.38 | -9.00 ± 30.75 | 0.2183 | -0.29 |
| **06** | **Accumulated anger/rage** | 81.37 ± | 65.37 ± | -16.00 ± | 0.0300 | -0.83 |

| | | | | | | |
|---|---|---|---|---|---|---|
| | | 18.73 | 22.43 | 19.34 | | |
| **07** | **Palpitations** | 70.37 ± 22.92 | 55.26 ± 20.31 | -15.11 ± 25.03 | 0.0647 | -0.60 |
| **08** | **Sleep disturbance** | 79.11 ± 20.82 | 63.84 ± 21.07 | -15.26 ± 27.41 | 0.0647 | -0.56 |
| **09** | **Headache/dizziness** | 73.00 ± 23.56 | 57.37 ± 22.51 | -15.63 ± 31.36 | 0.0723 | -0.50 |
| **10** | **Dry mouth** | 71.79 ± 23.71 | 59.18 ± 23.40 | -12.61 ± 29.76 | 0.1016 | -0.42 |
| **11** | **Loss of appetite** | 59.21 ± 22.75 | 46.97 ± 20.52 | -12.24 ± 21.39 | 0.0647 | -0.57 |
| **12** | **Fearfulness/startle response** | 69.79 ± 23.23 | 58.00 ± 22.61 | -11.79 ± 28.69 | 0.1040 | -0.41 |
| **13** | **Intrusive thoughts** | 82.90 ± 17.25 | 71.63 ± 17.00 | -11.26 ± 22.13 | 0.0723 | -0.51 |
| **14** | **Frequent sighing** | 80.37 ± 15.94 | 67.74 ± 23.43 | -12.63 ± 26.09 | 0.0737 | -0.48 |
| **15** | **Deep-seated resentment (Han)** | 76.90 ± 24.22 | 61.90 ± 24.98 | -15.00 ± 32.06 | 0.0769 | -0.47 |

**Note.** Values are presented as mean ± SD. p-values were calculated using paired t-tests.

**Table S7. Clinical scale changes from baseline to Week 4: per-protocol (PP) sensitivity analysis (n = 19).**

| Category | Variable | Baseline | Week 4 | Change | p-value (FDR) | Effect size |
|---|---|---|---|---|---|---|
| **HB Scales** | HB Personality Scale | 46.74 ± 8.86 | 39.26 ± 9.65 | -7.47 ± 12.66 | 0.0430 | -0.59[a] |
| | HB Symptom Scale | 46.21 ± 7.58 | 36.58 ± 9.68 | -9.63 ± 11.64 | 0.0120 | -0.83[a] |
| **Psychological Scales** | Depression (BDI-II) | 37.90 ± 11.00 | 24.47 ± 12.42 | -13.42 ± 13.55 | 0.0072 | -0.99[a] |
| | State Anxiety (STAI-S) | 45.68 ± 5.07 | 43.21 ± 4.98 | -2.47 ± 5.15 | 0.0759 | -0.48[a] |
| | Trait Anxiety (STAI-T) | 47.84 ± 5.73 | 42.90 ± 4.81 | -4.95 ± 5.86 | 0.0120 | -0.84[a] |
| | State Anger (STAXI-S) | 27.74 ± 7.78 | 20.90 ± 8.37 | -6.84 ± 8.69 | 0.0135 | -0.79[a] |
| | Trait Anger (STAXI-T) | 28.11 ± 7.16 | 21.90 ± 7.36 | -6.21 ± 9.08 | 0.0240 | -0.68[a] |
| | Anger Control | 19.84 ± 4.67 | 18.32 ± 4.18 | -1.53 ± 4.33 | 0.1958 | -0.35[a] |
| | Anger-Out | 18.16 ± 4.81 | 15.58 ± 5.16 | -2.58 ± 4.83 | 0.0596 | -0.53[a] |
| | Anger-In | 22.11 ± 5.11 | 19.21 ± 5.34 | -2.90 ± 5.72 | 0.0668 | -0.51[a] |
| | AAQ-II | 39.05 ± 10.65 | 32.32 ± 8.14 | -6.74 ± 10.22 | 0.0260 | -0.66[a] |
| **Quality of Life** | EQ-5D-5L Index | 0.68 ± 0.18 | 0.77 ± 0.10 | 0.09 ± 0.17 | 0.0596 | 0.49[b] |
| | EQ-5D-5L VAS | 51.32 ± 16.06 | 63.95 ± 13.50 | 12.63 ± 16.70 | 0.0144 | 0.76[a] |
| **HRV** | SDNN (ms) | 29.17 ± 14.34 | 29.06 ± 14.08 | -0.11 ± 16.68 | 0.5819 | 0.16[b] |
| | RMSSD (ms) | 25.22 ± 18.92 | 22.33 ± 10.72 | -2.89 ± 20.15 | 0.9621 | 0.01[b] |
| | PSI | 6.08 ± 0.65 | 6.04 ± 0.68 | -0.03 ± 0.62 | 0.9195 | -0.05[a] |
| | ln(TP) [$ms^2$] | 6.96 ± 0.57 | 6.82 ± 0.35 | -0.14 ± 0.62 | 0.5363 | 0.19[b] |
| | LF/HF | 1.37 ± 1.00 | 1.76 ± 1.57 | 0.39 ± 1.41 | 0.9195 | 0.04[b] |

**Note**. Values are presented as mean ± SD. [a], Effect size is Cohen's d for paired t-test. [b], Effect size is r for Wilcoxon signed-rank test.

**Table S8. Clinical scale and HB-VAS changes at follow-up (Week 8).**

Part A. Clinical Scale Changes (0W→8W and 4W→8W)

| Variable | Analysis | 0W→8W Change (Mean ± SD) | p-value (FDR) | 4W→8W Change (Mean ± SD) | p-value (FDR) |
|---|---|---|---|---|---|
| HB Personality | mITT (n=27) | -13.00 ± 9.88 | <0.001 | -2.78 ± 7.70 | 0.0589 |
| | PP (n=18) | -12.72 ± 10.20 | <0.001 | -3.72 ± 7.97 | 0.0741 |
| HB Symptom | mITT (n=27) | -16.11 ± 12.36 | <0.001 | -6.04 ± 9.05 | 0.0108 |
| | PP (n=18) | -17.44 ± 13.84 | <0.001 | -7.44 ± 8.66 | 0.0130 |
| Depression (BDI-II) | mITT (n=27) | -17.07 ± 15.19 | <0.001 | -3.63 ± 11.56 | 0.1236 |
| | PP (n=18) | -17.50 ± 17.25 | 0.0011 | -3.83 ± 12.91 | 0.2921 |
| State Anxiety | mITT (n=27) | -4.81 ± 4.73 | <0.001 | -3.04 ± 5.57 | 0.0286 |
| | PP (n=18) | -5.11 ± 5.52 | 0.0018 | -2.33 ± 6.37 | 0.2252 |
| Trait Anxiety | mITT (n=27) | -6.70 ± 6.31 | <0.001 | -1.63 ± 5.15 | 0.1236 |
| | PP (n=18) | -6.61 ± 6.21 | 0.0010 | -1.28 ± 5.31 | 0.3485 |
| State Anger | mITT (n=27) | -12.37 ± 9.83 | <0.001 | -4.52 ± 7.01 | 0.0108 |
| | PP (n=18) | -10.94 ± 11.02 | 0.0011 | -4.11 ± 8.04 | 0.0962 |
| Trait Anger | mITT (n=27) | -9.44 ± 8.28 | <0.001 | -2.52 ± 6.42 | 0.0747 |
| | PP (n=18) | -8.50 ± 8.28 | 0.0010 | -1.78 ± 6.32 | 0.2944 |
| AAQ-II | mITT (n=27) | -13.37 ± 9.69 | <0.001 | -6.04 ± 6.26 | <0.001 |
| | PP (n=18) | -13.56 ± 10.41 | <0.001 | -6.39 ± 6.38 | 0.0065 |
| EQ-5D Index | mITT (n=27) | 0.14 ± 0.21 | <0.001 | 0.05 ± 0.11 | 0.0390 |
| | PP (n=18) | 0.15 ± 0.23 | 0.0058 | 0.06 ± 0.11 | 0.0741 |
| EQ-5D VAS | mITT (n=27) | 18.30 ± 23.41 | <0.001 | 6.07 ± 15.09 | 0.0747 |
| | PP (n=18) | 20.83 ± 22.90 | 0.0019 | 10.28 ± 16.22 | 0.0676 |

Part B. HB-VAS 15-item Changes (0W→8W and 4W→8W)

| No | Symptom (mITT, n=27) | 0W→8W Change | p-value (FDR) | 4W→8W Change | p-value (FDR) |
|---|---|---|---|---|---|
| 01 | Chest tightness/breathlessness | -16.07 ± 20.17 | <0.001 | -4.19 ± 16.26 | 0.2065 |
| 02 | Rising sensation | -18.96 ± 22.08 | <0.001 | -6.85 ± 13.88 | 0.0373 |
| 03 | Heat sensation (face/chest) | -24.70 ± 24.69 | <0.001 | -12.33 ± 21.20 | 0.0195 |
| 04 | Lump sensation (throat/epigastrium) | -25.44 ± 24.34 | <0.001 | -10.22 ± 21.55 | 0.0373 |
| 05 | Feeling of injustice/resentment | -22.63 ± 25.53 | <0.001 | -8.37 ± 21.51 | 0.0715 |
| 06 | Accumulated anger/rage | -26.07 ± 25.00 | <0.001 | -9.67 ± 18.55 | 0.0373 |
| 07 | Palpitations | -25.89 ± 28.78 | <0.001 | -11.85 ± 23.66 | 0.0373 |
| 08 | Sleep disturbance | -16.67 ± 21.80 | <0.001 | -2.48 ± 18.04 | 0.2447 |
| 09 | Headache/dizziness | -27.78 ± 29.16 | <0.001 | -9.85 ± 21.09 | 0.0373 |

| | | | | | |
|---|---|---|---|---|---|
| **10** | **Dry mouth** | -23.37 ± 28.90 | <0.001 | -10.65 ± 22.29 | 0.0373 |
| **11** | **Loss of appetite** | -21.96 ± 28.21 | <0.001 | -7.98 ± 21.26 | 0.0715 |
| **12** | **Fearfulness/startle response** | -24.67 ± 26.85 | <0.001 | -9.93 ± 25.54 | 0.0715 |
| **13** | **Intrusive thoughts** | -21.26 ± 29.68 | 0.0010 | -10.44 ± 16.40 | 0.0142 |
| **14** | **Frequent sighing** | -28.15 ± 27.18 | <0.001 | -14.48 ± 19.94 | 0.0135 |
| **15** | **Deep-seated resentment (Han)** | -24.85 ± 30.47 | <0.001 | -8.00 ± 21.22 | 0.0715 |

**Table S9. HB screening status (HBSS ≥ 30) across assessment timepoints (mITT).**

Part A. Screening status at each time point

| Timepoint | N assessed | Screen-positive, n (%) | Screen-negative, n (%) |
|---|---|---|---|
| **0W (Baseline)** | 28 | 28 (100.0) | 0 (0.0) |
| **2W** | 27 | 25 (92.6) | 2 (7.4) |
| **4W (Post-treatment)** | 28 | 23 (82.1) | 5 (17.9) |
| **8W (Follow-up)** | 27 | 15 (55.6) | 12 (44.4) |

Part B. Conversion from baseline (0W) to Week 4

| Category | n (%) |
|---|---|
| **Screen-positive at 0W (N = 28)** | |
| **→ Converted to negative at 4W** | 5 (17.9) |
| **→ Remained positive at 4W** | 23 (82.1) |
| **Screen-negative at 0W (N = 0)** | |
| **→ New positive at 4W** | 0 (0.0) |

**Note**. McNemar's test not applicable at 0W vs. 4W as all participants were screen-positive at baseline.

**Table S10. Mixed model for repeated measures (MMRM) results for six primary clinical scales (mITT).**

Part A: Model fit indices and group variance

| Scale | Observations | Log-Likelihood | AIC | BIC | Group Var (SE) |
|---|---|---|---|---|---|
| **HB Symptom** | 82 | -284.28 | 576.56 | 586.18 | 36.90 (2.67) |
| **BDI-II** | 82 | -290.53 | 589.06 | 598.68 | 34.29 (2.42) |
| **STAI-S** | 82 | -236.56 | 481.12 | 490.75 | 6.86 (1.13) |
| **STAI-T** | 82 | -230.50 | 468.99 | 478.62 | 9.53 (1.36) |
| **STAXI-S** | 82 | -260.71 | 529.43 | 539.05 | 31.39 (2.84) |
| **STAXI-T** | 82 | -246.08 | 500.16 | 509.79 | 21.82 (2.38) |

Part B: Fixed effects estimates: coefficients and p-values

| Scale | Intercept (p) | Week 4 (p) | Week 8 (p) | Baseline (p) |
|---|---|---|---|---|
| **HB Symptom** | 28.720 (0.001) | -4.419 (0.015) | -10.391 (<0.001) | 0.276 (0.110) |
| **BDI-II** | 22.997 (<0.001) | -5.312 (0.009) | -8.752 (<0.001) | 0.204 (0.129) |
| **STAI-S** | 32.973 (<0.001) | 0.563 (0.595) | -2.494 (0.020) | 0.228 (0.130) |
| **STAI-T** | 30.558 (<0.001) | 0.263 (0.773) | -1.369 (0.139) | 0.259 (0.044) |
| **STAXI-S** | 15.643 (0.002) | -2.971 (0.017) | -7.307 (<0.001) | 0.291 (0.084) |
| **STAXI-T** | 15.749 (0.001) | -1.369 (0.185) | -3.934 (<0.001) | 0.288 (0.054) |

**Note**. Discrepancies between MMRM fixed-effect estimates and paired-test results (e.g., STAI-T and STAXI-T significant by paired test but not by MMRM at Week 4) reflect differences in model assumptions: paired tests compare observed means at each timepoint without adjusting for baseline or the covariance structure across visits, whereas MMRM estimates marginal change conditional on baseline and accounts for within-subject correlation across all timepoints simultaneously. These two approaches are complementary and not directly comparable.

**Table S11. Hwa-byung Symptom Scale exploratory responder analysis (mITT).**

| Timepoint | Scale | Criterion | N of analyzed | N of responders | Responder rate(%) |
|---|---|---|---|---|---|
| Week 4 | Hwa-byung Symptom Scale | ≥30% decrease from baseline | 28 | 8 | 28.6 |
| Week 8 | Hwa-byung Symptom Scale | ≥30% decrease from baseline | 27 | 16 | 59.3 |

**Note**. Responder criterion: ≥ 30% decrease from baseline HBSS. This threshold was not pre-specified and was adopted in the absence of an established MCID for the HBSS; findings are hypothesis-generating only.

**Table S12. Exploratory mediation path estimates and bootstrap indirect effect.**

Part A. Mediation Path Estimates and Statistical Associations

| Path | Description | Estimate (β/ρ) | Statistic / SD | p-value |
|---|---|---|---|---|
| **Path a** | ΔAAQ change from baseline | -7.036 | SD = 8.87 | 0.0011[†] |
| **Path c** | Total effect (Baseline HBSS → ΔHBSS) | -0.798 | β = -0.798 | 0.0007 |
| **Path b** | ΔAAQ → ΔHBSS (adj. Baseline HBSS) | 0.415 | β = 0.415 | 0.0410 |
| **Path c'** | Direct effect (Baseline HBSS → ΔHBSS \| ΔAAQ) | -0.595 | β = -0.595 | 0.0105 |
| **Correlation** | Spearman ρ (ΔAAQ vs. ΔHBSS) | 0.532 | ρ = 0.532 | 0.0035 |
| **Model Fit** | $R^2$ (Full mediation model) | 0.463 | - | - |

**Note**. [†], Calculated using Wilcoxon signed-rank test (W = 41.5).

Part B. Bootstrap Indirect Effect Analysis

| Effect | Mean Estimate | Bootstrap 95% CI (Lower) | Bootstrap 95% CI (Upper) | Significant |
|---|---|---|---|---|
| Indirect effect | -3.0156 | -6.4187 | -0.4030 | Yes |

**Note**. Bootstrap results based on 5,000 resamples (seed = 42). Significance is determined by the 95% CI excluding zero. All mediation findings are exploratory and should be interpreted with caution given the single-arm pre-post design; causal inference is not warranted.

Part C. Exploratory Logistic Regression for Clinical Outcome

| Model | Independent Var. | Odds Ratio | p-value | N |
|---|---|---|---|---|
| HB Screening Status (4W) | ΔAAQ | 1.226 | 0.0286 | 28 |

**Note**. Model adjusted for baseline HBSS. Outcome defined as screen-positive status at Week 4.

**Table S13. Adherence–outcome correlation: Spearman's ρ between app use days (minimum and maximum) and change in clinical outcomes from baseline to Week 4 (mITT).**

| Outcome (Δ Baseline to 4W) | Adherence Metric | Spearman's ρ | p (raw) | p (FDR) |
|---|---|---|---|---|
| HB Symptom Scale | Min use days (app launch) | 0.025 | .898 | .982 |
| | Max use days (full session) | 0.149 | .450 | .982 |
| Depression (BDI-II) | Min use days (app launch) | -0.087 | .661 | .982 |
| | Max use days (full session) | 0.004 | .982 | .982 |
| State Anxiety (STAI-S) | Min use days (app launch) | -0.202 | .303 | .982 |
| | Max use days (full session) | -0.054 | .786 | .982 |
| Trait Anxiety (STAI-T) | Min use days (app launch) | 0.056 | .776 | .982 |
| | Max use days (full session) | 0.196 | .318 | .982 |
| State Anger (STAXI-S) | Min use days (app launch) | 0.059 | .767 | .982 |
| | Max use days (full session) | 0.122 | .535 | .982 |
| Trait Anger (STAXI-T) | Min use days (app launch) | 0.018 | .926 | .982 |
| | Max use days (full session) | 0.103 | .603 | .982 |
| Acceptance (AAQ-II) | Min use days (app launch) | -0.013 | .947 | .982 |
| | Max use days (full session) | 0.112 | .571 | .982 |

**Note**. Spearman's rank correlation coefficient (ρ) was utilized to analyze the relationship between adherence and clinical outcomes in the modified intention-to-treat (mITT) population (N = 28). Adherence was operationalized as the number of days with app usage, ranging from 'Minimum' (days with at least one app launch) to 'Maximum' (days with at least one full diaphragmatic breathing session completion). Benjamini-Hochberg FDR correction was applied across all 14 correlation pairs, and no statistically significant associations were observed ($p < .05$). All p-FDR values resulted in approximately .982, indicating that the magnitude of clinical improvement was independent of the frequency of app usage or session completion in this study.
